# A Synbiotic Infant Formula Promotes Balanced Immune-Microbiome Interactions in Term Infants: A Randomized Controlled Trial

**DOI:** 10.64898/2026.10.05.26364773

**Authors:** Hugo Barcenilla, Cheong Kwong Chung, Maria Rosario Z. Capeding, Mitzi Trinidad-Aseron, Vinna Marie R. Tenorio-Quiñones, Anna James, Qi Chen, Laura Gonzalez, Anette Johnsson, Noura Darwish, Haolin Li, Jacob Agerbo Rasmussen, Janne Marie Moll, William Stauch, Jochen Schwenk, Tadepally Lakshmikanth, Norbert Sprenger, Nicholas P. Hays, Mario Noti, Petter Brodin

**Author notes:** **Correspondence:** Professor Petter Brodin, M.D., Ph.D. These authors contributed equally and share the first authorship (Hugo Barcenilla, Cheong Kwong Chung, Maria Rosario Z. Capeding, Mitzi Trinidad-Aseron, Vinna Marie R. Tenorio-Quiñones).

## Abstract

Formula-fed infants have altered gut microbiota and immune development relative to breastfed infants. Human milk oligosaccharides (HMOs) and microbes able to ferment these, like *Bifidobacterium infantis,* promote healthy immune-microbiome interactions in breastfed infants. In this randomized trial, 240 healthy term infants in the Philippines received a synbiotic formula (SynF; seven HMOs, bovine milk oligosaccharides, and B. infantis LMG11588) or control formula (CtrlF) for 6 months. SynF supported growth non-inferior to CtrlF and near universal *B. infantis* colonization (*vs* <10% in CtrlF). Markers of intestinal inflammation were lower in children fed SynF while secretory IgA was higher. SynF fed infants showed lower markers of systemic inflammation in plasma, slower immune cell maturation, and fewer upper respiratory tract infections (URTI) during their first 6 months of life. These findings suggest that synbiotic formula can dampen inflammatory responses to colonizing microbes and mild infections in fully formula-fed, term infants. Trial registration: NCT06073652.

## Main

Breastfeeding is the gold standard for infant nutrition, providing substantial benefits that lead to a balanced development of the microbiota and immune system via early life immune-microbiome interactions that promote long-term health^1^. Breast milk provides essential nutrients to the child but also a complex array of bioactive components, including human milk oligosaccharides (HMOs), lactoferrin^2^ and other bioactive proteins (e.g. immunoglobulins)^3^, as well as a diverse microbial community that collectively shape the infant gut microbiome and limit inflammatory responses to commensal microbes^4,5^. Breastfed infants harbor gut microbiomes dominated by infant-type *Bifidobacterium* species, particularly *Bifidobacterium longum* subsp. *infantis*, which has co-evolved with the mother-infant dyad to efficiently metabolize HMOs^6–8^. This bifidobacteria-dominant microbiome has been associated with reduced markers of gut inflammation, enhanced mucosal immune defenses upon infection and lower risks of allergic diseases, and metabolic disorders^8,9^. However, despite strong recommendations for exclusive breastfeeding during the first six months of life, global breastfeeding rates remain suboptimal, with 30–40% of infants receiving infant formula either exclusively or as a supplement^1^.

Formula-fed infants exhibit distinct gut microbiome profiles characterized by lower *Bifidobacterium* abundance, higher microbial diversity, and increased prevalence of potentially pathogenic taxa such as *Clostridioides difficile* and *Ruminococcus gnavus*^10,11^. This altered microbiome composition has been associated with elevated gut inflammation, increased intestinal permeability, and a more activated systemic immune phenotype^12,13^. A finding of particular concern is that recent studies have documented a generational decline in *B. infantis* colonization among infants in industrialized nations, which has been proposed as one potential contributor to the rising incidence of immune-mediated allergic and autoimmune diseases^8^. These observations have motivated efforts to develop infant formulas supplemented with bioactive components that can narrow the gap between formula-fed and breastfed infants.

Human milk oligosaccharides represent the third largest solid component of human milk, with concentrations ranging from 5–15 g/L^5,14^. These structurally diverse complex carbohydrates are indigestible by the infant and instead serve as selective substrates for beneficial gut bacteria, particularly *B. infantis*, which possess specialized enzymatic machinery to metabolize HMOs^6,15^.

Beyond their prebiotic function, HMOs exert direct immunomodulatory effects, serving as decoy receptors that prevent pathogen adhesion to epithelial cells and modulating immune cell responses^16,17^. Advances in biotechnology have enabled the large-scale production of HMOs identical to those found in human milk, including 2’-fucosyllactose (2’FL), lacto-N-tetraose (LNT), lacto-N-neotetraose (LNnT), and sialylated oligosaccharides^18^. Clinical trials have demonstrated that infant formulas supplemented with one to six manufactured HMOs are safe, well-tolerated, and support normal growth, with some studies reporting stool microbiome shifts towards a more breastfed-like microbial composition and even reduced morbidity^19–22^.

Similarly, bovine milk-derived oligosaccharides (MOS or galacto-oligosaccharides) and probiotic *Bifidobacterium* strains have been evaluated as formula supplements^23^. MOS at concentrations up to 7.2 g/L have been shown to promote softer stools, increased fecal bifidobacteria abundances, and a shift in overall microbiota composition closer to that of breastfed infants^24^. Supplementation with *B. infantis* has demonstrated safety and tolerability in multiple trials, with evidence of persistent colonization, increased short-chain fatty acid production^25–27^, and modulation of fecal cytokine profiles suggestive of reduced enteric inflammation^8^. Although several studies have assessed probiotics, particularly supplementation with *B. infantis*, most of these have focused on additive effects in breastfed infants. It remains unknown whether synbiotic supplements benefit exclusively formula-fed infants, and no prior study has characterized local immune-microbiome interactions and systemic immune development in relation to nutritional interventions so early in life.

Here, we report results from a randomized, double-blind, controlled trial evaluating an infant formula supplemented with a synbiotic blend comprising seven human identical manufactured HMOs (2.64 g/L total), bovine milk oligosaccharides (4.56 g/L), and *Bifidobacterium longum* subsp. *infantis* LMG11588 (10⁶ CFU/g) in term-born, formula-fed infants. The HMO concentrations in this formula fall within the range observed in mature human milk^12^. The primary objective was to demonstrate non-inferior growth compared with a standard infant formula at 4 months of age. Predefined secondary objectives included assessment of safety, gastrointestinal tolerance, gut microbiome composition, and fecal biomarkers of gut health. In a pre-specified sub-study, we performed comprehensive systemic immune profiling using mass cytometry, multidimensional plasma protein profiling, and vaccine response assays to characterize how synbiotic supplementation affects immune-microbiome interactions and systemic immune maturation during the first six months after birth. We further examined associations between gut microbiome developmental trajectories, stool metabolites, and systemic immune markers. Our findings demonstrate that synbiotic formula promotes a coordinated maturation of the gut-microbiome with a dominance of bifidobacteria, reduced markers of inflammation in the gut and in systemic circulation, and fewer and shorter episodes of symptomatic respiratory infections. Together, these outcomes are consistent with a real-world illustration of a more balanced immune states and a more regulated inflammatory set point early in life.

## Results

### Study infants and baseline characteristics

A total of 243 infants in Manila, the Philippines were screened and 240 were randomized into the CtrlF (n=122) and SynF (n=118) groups forming the Intention-To-Treat (ITT) dataset (**Fig. 1a**). Seventeen infants were excluded from the Full Analysis Set (FAS) as they did not receive any study product, leaving 114 and 109 in the CtrlF and SynF groups, respectively. After excluding infants with major protocol deviations, 113 in the CtrlF group and 108 in the SynF group comprised the Per-Protocol (PP) population (**Fig. 1a**). For the blood sample analysis, N=43 and N=42 infants representative of CtrlF and SynF infants respectively were selected as Biomarker Analysis Set (BAS) (Fig. 1a). One additional SynF infant, sampled at baseline but not part of the FAS, contributes to the baseline blood measurements only (n=43 SynF infants at baseline in Fig. 5c and 5f). A total of 217 (90%) enrolled infants completed the study. Infant baseline demographic and household characteristics for the ITT and BAS were similar for SynF and CtrlF groups (**Fig. 1b**). These are consistent with the Full-Analysis Set (FAS) population and patients were monitored for 6 months and blood sampled at baseline, and 2- and 4-months while stool was collected at baseline, 2-, 4- and 6 months of life (**Fig. 1c**).

**Figure 1.**
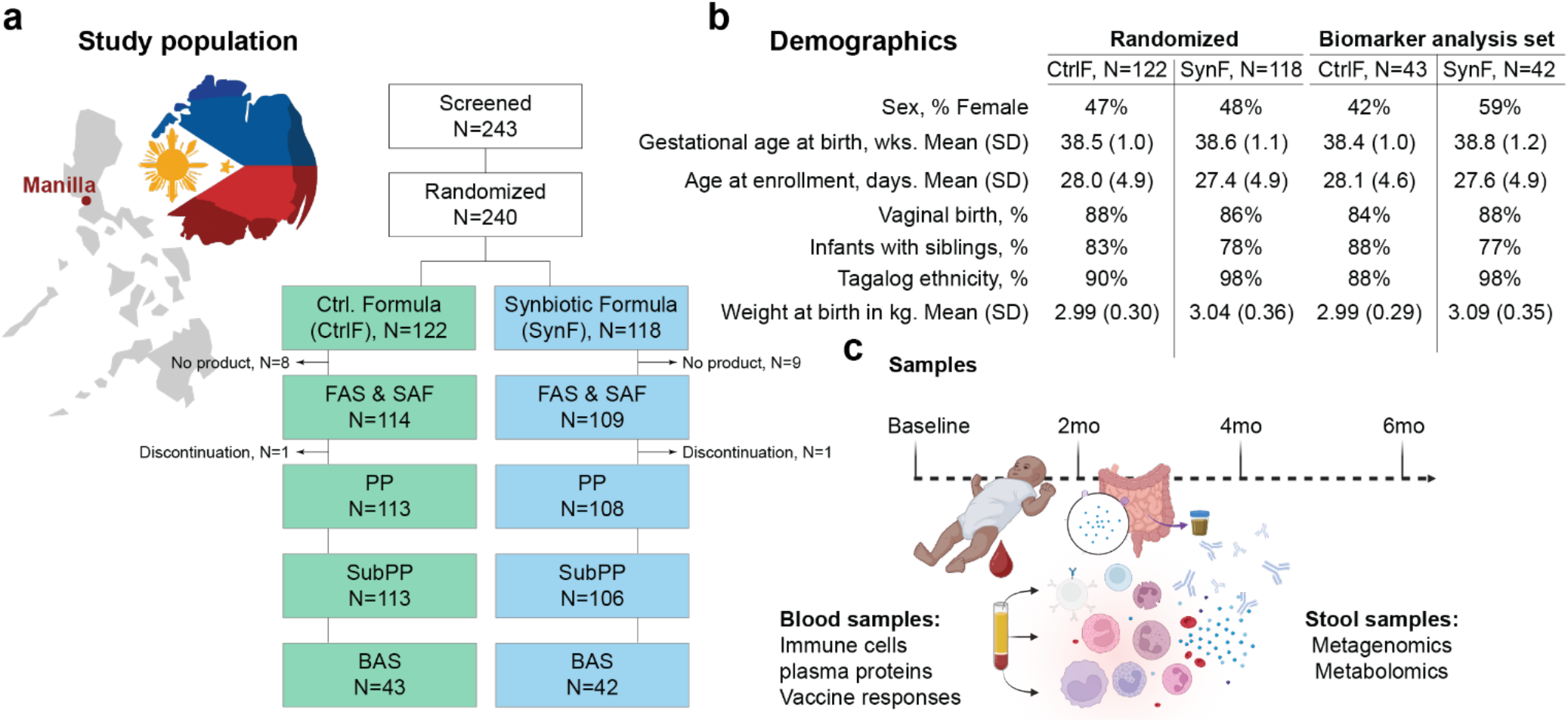
Study design and cohort. a) Infants born in Manila, Philippines randomized to Synbiotic formula (SynF), or Control Formula (CtrlF). Patient randomization and follow-up. FAS: Full analysis set; SAF: Safety analysis set; PP: Per protocol; SubPP: Sub-Per-Protocol; BAS: Biomarker analysis set. b) Baseline demographics in SynF and CtrlF groups. Neither of the average measurements significantly differed between these groups. c) Stool and blood sampling for systems-level profiling in biomarker analysis set of participants.

### Anthropometrics

Weight gain velocity was non-inferior in the SynF group compared to the CtrlF group in the FAS and PP populations with a mean difference (95% CI) of 1.01 (−0.67, 2.68) (p<0.001). Weight gain velocity mean from baseline to 4 months was (95% CI) 28.0 (26.7, 29.3) and 27.0 (25.7, 28.3) g/day for the SynF (n=107) and the CtrlF (n=110) groups, respectively, in both FAS and PP datasets. No significant differences were observed between groups in weight-for-age, length-for-age, weight-for-length, or head circumference-for-age *z*-scores at any time point and all were within normal range (**Fig. 2a-d**).

**Figure 2.**
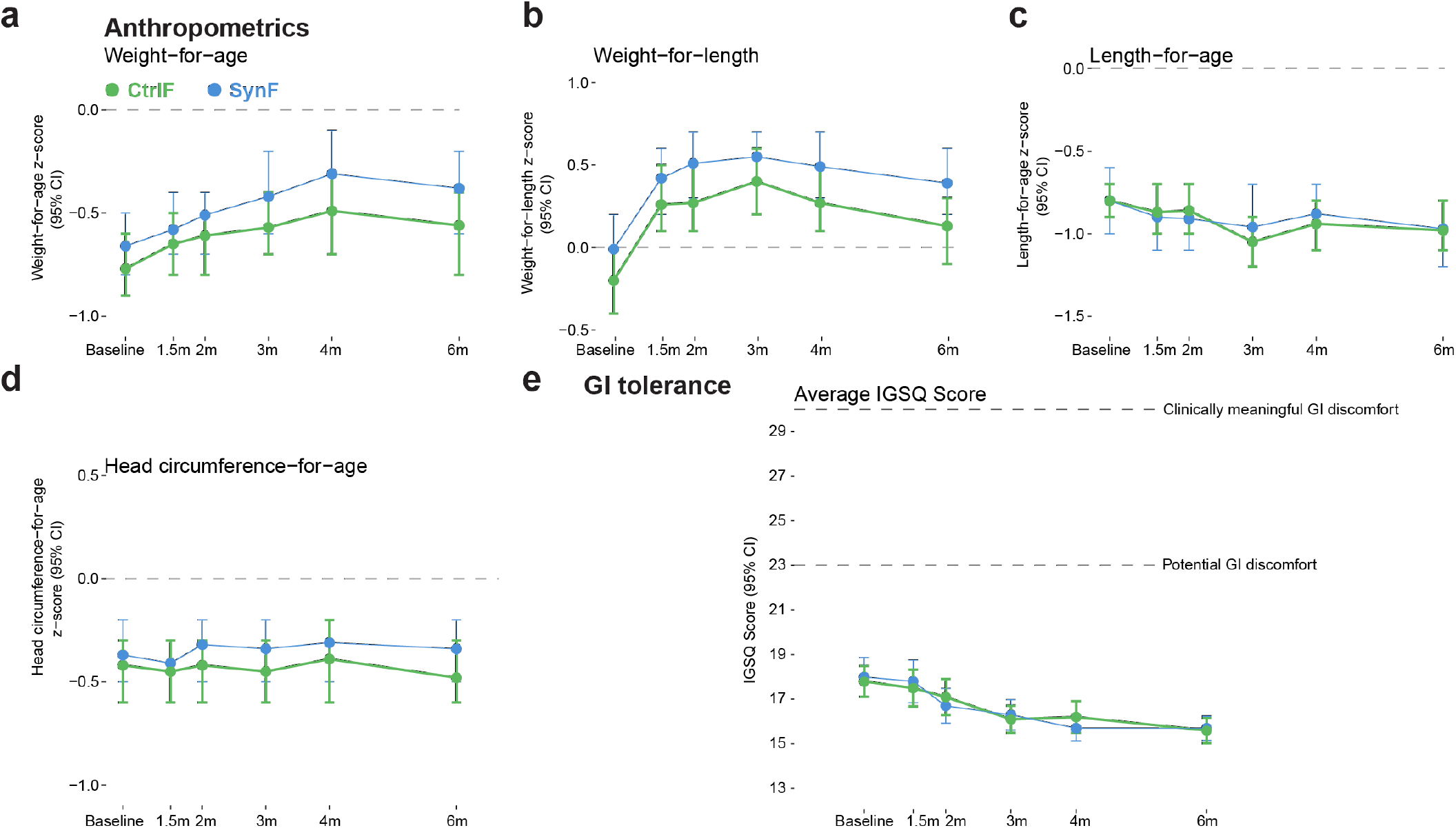
Growth and gastrointestinal discomfort. a) Infants fed Synbiotic Formula (SynF), or Control Formula (CtrlF), monitored for Weight-for-age, b) Weight-for-length, c) Length-for-age, and d) head circumference-for-age. e) Gastrointestinal, GI tolerance measured by IGSQ-13 forms answered by parents or another responsible caretaker.

### Gastrointestinal tolerance

Any nutritional intervention can be associated with intestinal discomfort in young children. We assessed Infant Gastrointestinal Symptom Questionnaire, IGSQ scores^28^ and mean levels were low (≤18.0) in both groups over time, indicating good GI tolerance (**Fig. 2e**). Consistent with these findings, parent-reported stooling patterns from the 3-day diary showed that stools were slightly more frequent among SynF fed infants compared with CtrlF fed infants at months 3 (mean 2.0 *vs* 1.6; p=0.0457), 4 (mean 1.9 *vs* 1.4; p=0.0014), and 6 (mean 1.8 *vs* 1.5; p=0.0254). Stools were also generally softer among SynF fed infants at months 1.5 (1.8 *vs* 2.0; p=0.0002), 2 (1.8 *vs* 2; p=0.0006), and 4 (1.8 *vs* 2; p=0.0013) compared to CtrlF fed infants though mean consistency in both groups was ‘mushy-soft.’ The number of days on which infants had difficulty passing stools was low and similar in both groups (all p>0.05).

### Safety and exploratory efficacy

Adverse Events, AE incidence was similar in both groups (51.4% and 53.5% in SynF and CtrlF, respectively) (**Table 1**). Eight Serious Adverse Events, SAEs were reported (4 events in 4 infants per group), including one fatal event unrelated to the study in the CtrlF group (aspiration pneumonia). The number of infection-related AEs was similar in both groups (n=43 in SynF and n=48 in CtrlF). Fewer URTI events were reported among infants in the SynF group compared to those in the CtrlF group (19 vs 41 events of the MedDRA term upper respiratory tract infection, respectively; p=0.009; 25 vs 46 when viral URTI is included, p=0.023) (**Table 2**).

**Table 1.** Adverse events summary.

|  | **SynF,** (n=109) |  | **CtrlF,** (n=114) |  |
|---|---|---|---|---|
|  | **Events,** n | **Infants,** N (%) | **Events,** n | **Infants,** N (%) |
| Any AE | 79 | 56 (51.4%) | 93 | 61 (53.5%) |
| Serious AE | 4 | 4 (3.7%) | 4 | 4 (3.5%) |
| Severity |  |  |  |  |
| Mild | 71 | 53 (48.6%) | 87 | 60 (52.6%) |
| Moderate | 5 | 5 (4.6%) | 3 | 3 (2.6%) |
| Severe | 3 | 3 (2.8%) | 3 | 3 (2.6%) |
| Relation |  |  |  |  |
| Unlikely | 2 | 1 (0.9%) | 1 | 1 (0.9%) |
| Unrelated | 77 | 55 (50.5%) | 92 | 60 (52.6%) |

**Table 2.** Infections and infestations as adverse events.

|  | **SynF** (n=109) |  | **CtrlF,** (n=114) |  |
|---|---|---|---|---|
|  | **Events,** n | **Infants,** N (%) | **Events,** n | **Infants,** N (%) |
| **Infections and infestations** | 56 | 43 (39.4) | 71 | 48 (42.1) |
| Acrodermatitis | 1 | 1 (0.9) | 1 | 1 (0.9) |
| Acute sinusitis | 1 | 1 (0.9) | 0 | 0 (0) |
| Bronchitis | 7 | 7 (6.4) | 4 | 3 (2.6) |
| Carbuncle | 1 | 1 (0.9) | 0 | 0 (0) |
| Exanthema subitum | 1 | 1 (0.9) | 0 | 0 (0) |
| Gastroenteritis | 6 | 4 (3.7) | 0 | 0 (0) |
| Gastroenteritis viral | 0 | 0 (0) | 1 | 1 (0.9) |
| Nasopharyngitis | 3 | 3 (2.8) | 4 | 4 (3.5) |
| Pneumonia | 8 | 8 (7.3) | 10 | 9 (7.9) |
| Pneumonia aspiration | 0 | 0 (0) | 1 | 1 (0.9) |
| Respiratory tract infection | 3 | 3 (2.8) | 2 | 2 (1.8) |
| Upper respiratory tract infection | 19 | 18 (16.5) | 41 | 28 (24.6) |
| Urinary tract infection | 0 | 0 (0) | 1 | 1 (0.9) |
| Viral pharyngitis | 0 | 0 (0) | 1 | 1 (0.9) |
| Viral upper respiratory tract infection | 6 | 4 (3.7%) | 5 | 5 (4.4) |

### Intestinal barrier and immune biomarkers

SynF compared to CtrlF formula feeding was associated with reduced inflammatory biomarkers in stool samples, including lower levels of calprotectin at 4 months, and alpha-1-antitrypsin, AAT at 2, 4, and 6 months (all p<0.05) (**Fig. 3a-b**). This indicates a less inflammatory response to colonizing microbes in SynF fed infants. Secretory IgA, sIgA in stool reflects an intestinal immune system that is engaging with colonizing microbiota in a controlled, non-inflammatory way, rather than one that is either immunologically naive or actively inflamed^29^. In breastfed infants, sIgA is provided via breastmilk and partly explains the protective effect of breastmilk from the catastrophic intestinal inflammation in necrotizing enterocolitis, NEC in preterms^30^. In formula-fed infants, all sIgA is produced by the host and translocated via the polymeric Ig receptor, pIgR^29^. At 4 months, the level of sIgA was higher in SynF *vs* CtrlF infants (p=0.0341) (**Fig. 3c**). Another biomarker of immune-microbiome interactions early in life is fecal pH which reflect the fermentation chemistry of gut microbes with lower pH in breastfed than formula fed infants^31^ and among breastfed infants, pH is lower when the relative abundance of Bifidobacteria is higher^32^. We found that fecal pH was consistently lower in SynF compared to the CtrlF group from 2-6 months (p<0.0001) as another example of the synbiotic formula providing a more breastfed-like state as compared to control formula (**Fig. 3d**).

**Figure 3.**
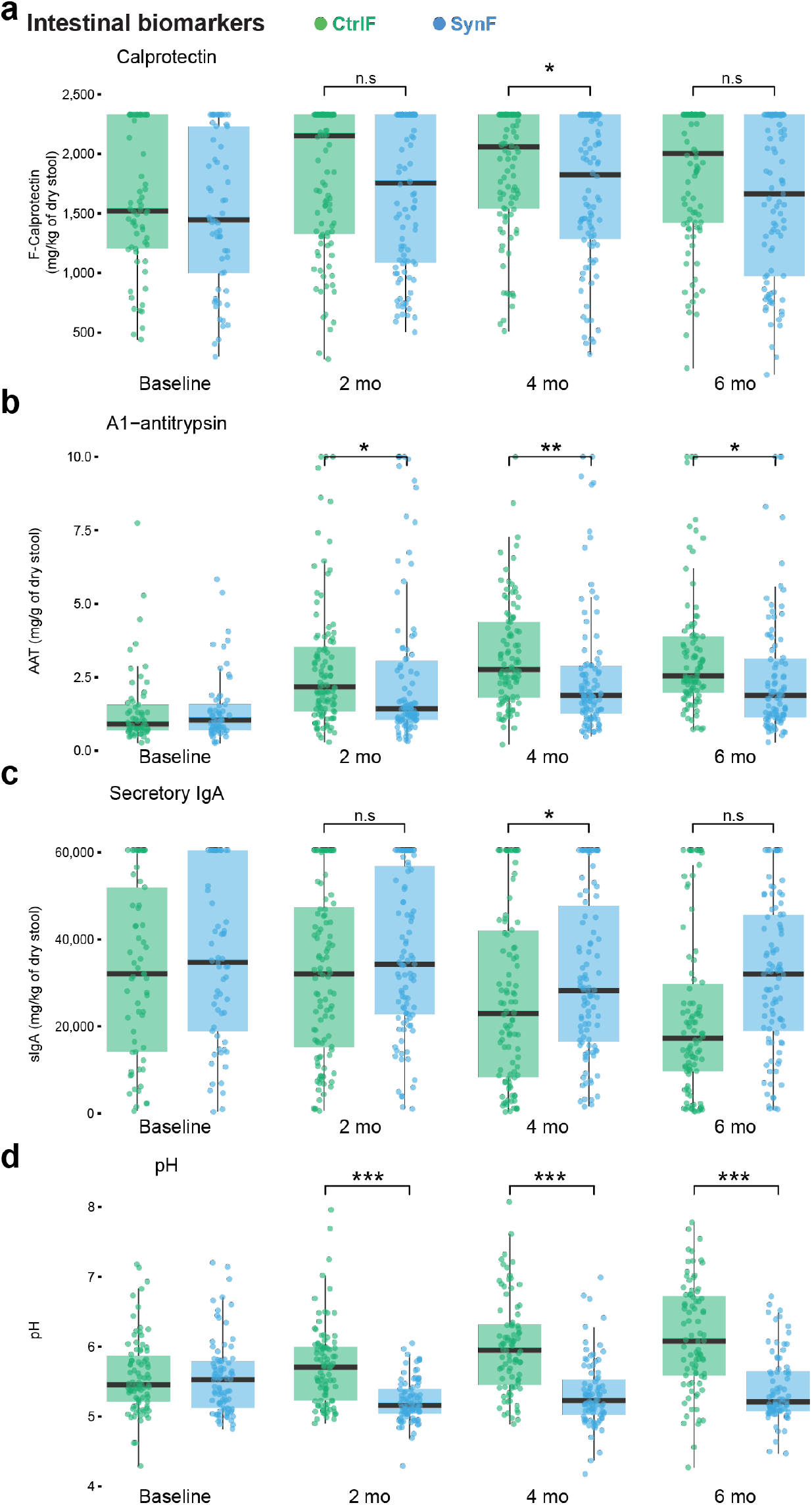
Markers of intestinal health in formula-fed infants. Infants fed Synbiotic formula (SynF) in blue, or Control Formula (CtrlF) in green analyzed for a) fecal Calprotectin, b) fecal A1-Antitrypsin, c) secretory IgA in stool and d) fecal pH at the indicated time points. n.s= not significant, * p<0.05, ** p<0.01, *** p<0.001.

### Gut microbiome development and metabolites

Microbiome diversity was assessed using alpha-diversity metrics (Richness, Shannon index, and Faith’s phylogenetic diversity [Faith’s PD]) and beta-diversity metrics (Bray-Curtis dissimilarity and weighted UniFrac distance) and differed between SynF and CtrlF groups (p<0.0001) from 2-6 months (**Fig. 4a**). Low intra-individual (alpha-) diversity is generally observed in breastfed infants because of the dominance of Bifidobacterium species able to ferment HMOs. We measured bifidobacterium species by metagenomic sequencing and found higher abundances (mainly *B. infantis, B. breve, B. longum, B. bifidum*) from 2-6 months of age, both in relative (data not shown) and absolute terms (**Fig. 4b**). *B. infantis* dominated in SynF fed infants and showed the strongest increase over time (p<0.0001) (**Fig. 4b**). From 2 months, *B. infantis* (probiotic LMG11588 and endogenous strains) prevalences were virtually 100% in SynF fed infants, while most CtrlF fed infants had barely detectable *B. infantis* through their first 6 months (**Fig. 4b**). Infants born via C-section and without older siblings showed the strongest bifidobacteria increase in SynF compared to CtrlF fed infants from 2-6 months (**Supplementary Fig. 1a-b**). As expected from the strong increase in infant-type bifidobacteria, an increase in internal HMO utilization capacity is seen in the SynF group from 2 to 6 months: GH95, GH29 for fucosylated HMOs; GH33 for sialylated HMOs; GH42, GH112 for type-1 core structures and GH20 (**Fig. 4c**). The CtrlF formula fed infants showed higher levels of secreted GH95, GH29, GH136 and GH2 at 6 months. The SynF group is characterized by high relative (and absolute) abundance of infant type bifidobacteria, the CtrlF group has higher relative abundance of other taxa that generally colonize to higher abundances, later, after 6 months. Among those, the mucosal glycan forager *Ruminococcus gnavus* was higher in CtrlF *vs* SynF fed infants from 2-6 months (**Fig. 4b**). All in all, these results show that feeding synbiotic formula containing HMO, BMO, and probiotic *Bifidobacterium infantis* strongly impacts gut microbiome development in favor of the probiotic strain and other HMO metabolizing microbes commonly seen in breastfed infants.

**Figure 4.**
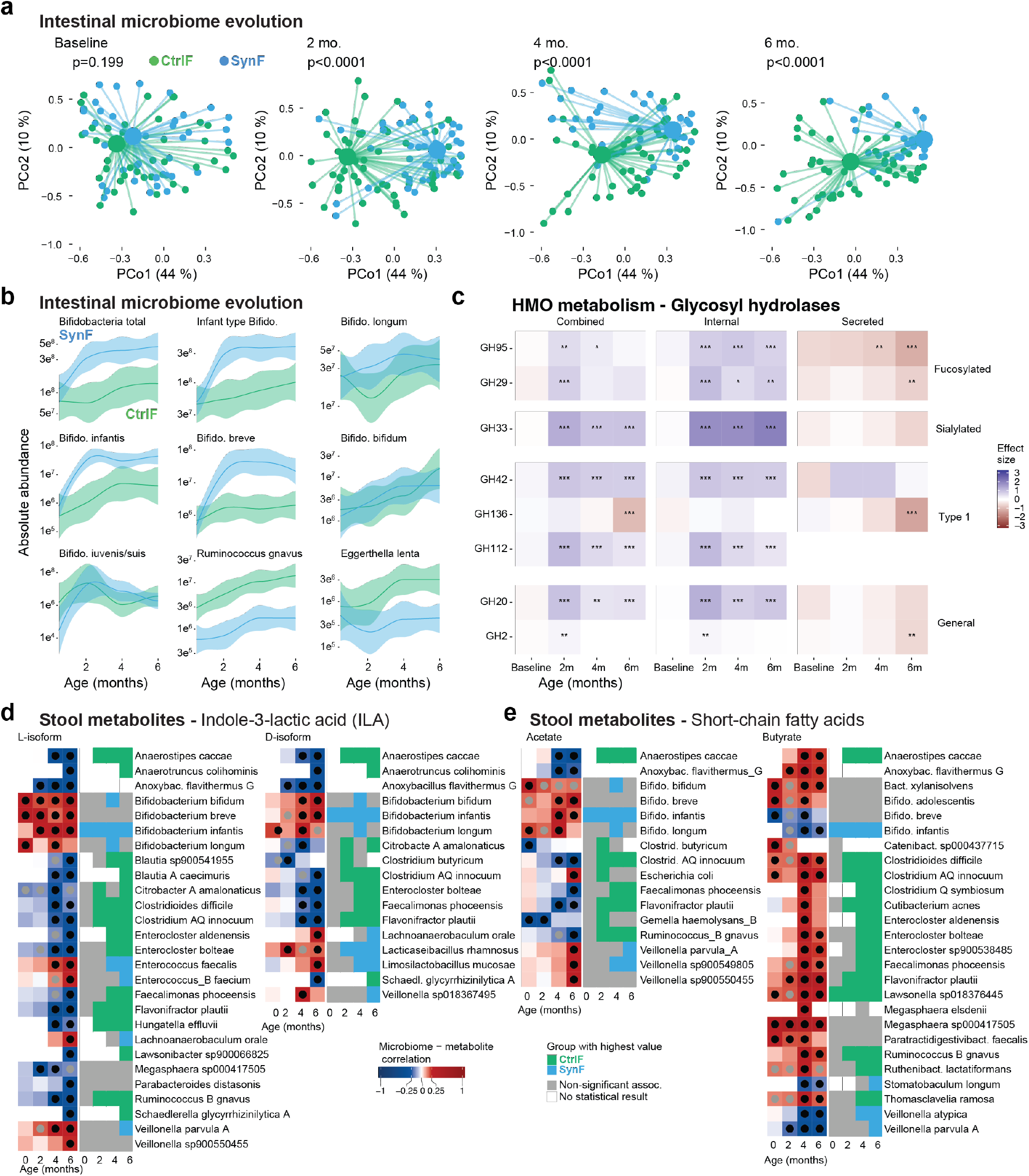
Microbiome evolution and metabolite profiles impacted by formula type. Infants fed Synbiotic formula (SynF) are shown in blue and infants fed Control formula (CtrlF) in green. **a**) Principal coordinates analysis (PCoA) of weighted UniFrac distances from fecal metagenomic sequencing at baseline, 2, 4 and 6 months; p-values compare the CtrlF and SynF groups at each timepoint. **b**) Absolute abundances of the indicated Bifidobacterium and other bacterial species over time in the SynF and CtrlF groups, shown as mean ± CI. **c**) Effect sizes for the difference in abundance of HMO-metabolizing glycosyl hydrolase (GH) gene programs between groups (combined, internal and secreted enzymes), grouped by HMO substrate (fucosylated, sialylated, type 1 and general); positive values (blue) indicate higher abundance in SynF and negative values (red) higher abundance in CtrlF. Stars indicate significance * p<0.05, ** p<0.01, *** p<0.001, **** p<0.0001. **d**) Stool indole-3-lactic acid (ILA; L- and D-isoforms), and **e**) short-chain fatty acids (acetate and butyrate), measured at the same timepoints as the metagenomic sequencing by targeted mass spectrometry. Heatmaps show the correlation coefficient between the indicated microbe and each metabolite at 0, 2, 4 and 6 months (dots mark significant correlations); the adjacent panels show the group with the highest value for each microbe (CtrlF, green; SynF, blue; grey, non-significant association; white, no statistical result).

### Microbial metabolites in relation to formula feeding

Metabolites produced by microbes are known to exert immunomodulatory effects and contribute to the establishment of healthy immune-microbiome interactions early in life^33^. For example, we previously found that indole-3 lactic acid (ILA), a tryptophan metabolite was associated with lower levels of intestinal inflammation and immunomodulation in breastfed infants fed probiotic *B. infantis*^8^. In the formula-fed infants studied here we also found that ILA in stool was positively correlated with *B. infantis* and *B. bifidum* abundances and strongly associated with SynF feeding between 2-6 months (**Fig. 4d**). Short chain fatty acids (SCFAs) are another class of metabolites known to exert beneficial effects on epithelial cells and developing immune cells early in life^33^. SCFA acetate is associated with breastfeeding and the presence of HMO metabolizing bifidobacteria^34^, and acetate has been associated with reduced risks of allergy, asthma, and atopy^35^. In SynF-fed infants we measured high acetate concentrations from 2 to 6 months, like ILA (**Fig. 4d**). Butyrate has also been associated with reduced risks of immune mediated disease, but butyrate is usually a product of clostridial colonization later in infancy and in SynF children we found an inverse correlation between butyrate and *B. infantis* abundances (**Fig. 4d**). A similar pattern was seen for propionate levels (**Supplementary Fig. 2**). These findings indicate that synbiotic formula retains a developing microbiome in a *B. infantis,* acetate and ILA dominated state, characteristic of the intestinal milieu of exclusively breastfed^8^ and associated with favorable immune-microbiome interactions in contrast to the accelerated maturation upon early weaning (<3 months) associated with increased risks of immune-mediated disease^36^.

### Systemic immune profile development

To directly assess the effects of CtrlF *vs* SynF on immune development we performed systems-level analyses of longitudinal whole blood samples collected at baseline, and at 2- and 4 months within the BAS population. Mass cytometry analysis of 21,865,760 whole blood cells was manually classified by known marker combinations into 27 core immune cell populations (**Supplementary Fig. 3**). Principal component analysis of immune cell frequencies showed that overall immune cell composition is structured by age (**Fig. 5a, b**), and that the cells increasing after birth are B cells, neutrophils, and CD8^+^ T effector cells, while the cells decreasing in blood after birth are classical monocytes, regulatory T cells, and central memory CD4^+^ T cells^37^, possibly reflecting gut homing of these cells^8^ (**Supplementary Fig. 4**). Using pseudotime^38^ derived from the top time-varying cell populations as an integrated readout of immune cell maturation, SynF and CtrlF infants were indistinguishable at baseline (d = 0.12, p = 0.943) and at 2 months (d = −0.08, p = 0.628), but trended lower in SynF by 4 months (d = −0.46, p = 0.057) suggestive of a slightly slower maturation of immune cells in blood in children fed synbiotic formula (**Fig. 5c**).

**Figure 5.**
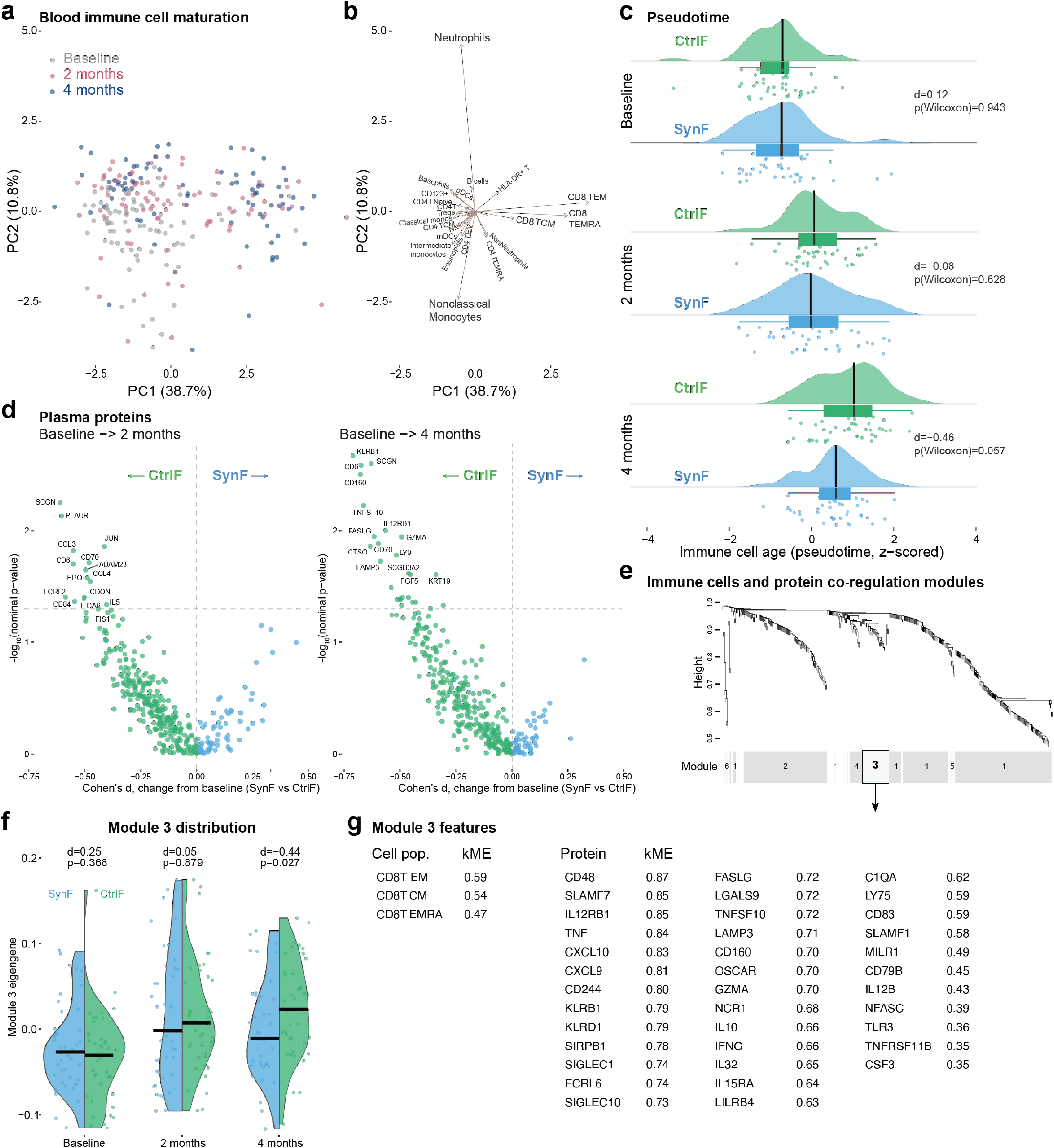
Immune development in relation to feeding practices. Infants fed the synbiotic formula (SynF) or control formula (CtrlF). (**a**) Principal component analysis (PCA) of 27 manually gated CyTOF immune cell population frequencies (percent of CD45+ cells), colored by timepoint (baseline, 2 months, 4 months); PC1 and PC2 explain 38.6% and 10.8% of variance, respectively. (**b**) The same PCA shown as a biplot, with loading vectors indicating which cell populations drive each principal component (neutrophils and nonclassical monocytes load on PC2; CD8 memory/effector subsets — central memory, effector memory and terminally differentiated effector memory — load on PC1). (**c**) Immune maturation age (pseudotime, z-scored, derived from the top time-varying cell populations) by feeding group at each timepoint; raincloud plots show distributions for CtrlF (green) and SynF (blue), with Cohen’s d and Wilcoxon p-values for the cross-sectional comparison at each timepoint. (**d**) Change from baseline (Cohen’s d, x-axis) versus nominal significance (−log10 p-value, y-axis) for each of 358 Olink-measured plasma proteins, comparing SynF with CtrlF, at 2 months (left) and 4 months (right); points are colored by which feeding group showed the larger change from baseline (CtrlF, green; SynF, blue), the 15 most differentially regulated proteins per panel are labelled, and the dashed horizontal line marks nominal p = 0.05. (**e**) Weighted gene co-expression network analysis (WGCNA) combining 27 CyTOF population frequencies and 358 plasma proteins into one joint co-regulation network; the dendrogram shows hierarchical clustering by topological overlap, with modules identified by dynamic tree cut and numbered by size (Module 1 largest, 203 features; Module 6 smallest, 6 features). (**f**) Module 3, a 40-feature CD8+ T effector/memory axis (3 cell populations and 37 proteins): eigengene by feeding group at each timepoint (individual samples as points, black bars mark group medians); Cohen’s d and nominal Wilcoxon p-values for SynF versus CtrlF are shown per timepoint. (**g**) Cell populations and proteins in Module 3, ranked by module membership (kME, correlation of each feature with the module eigengene).

Plasma protein profiling^39^ (Olink Explore 384 Inflammation I) revealed a reproducible pattern in which markers of immune activation and chemotaxis, including KLRB1, CD160, TNFSF10, FASLG, GZMA, IL12RB1, CD70, CD6, LY9, LAMP3, CTSO, CCL3, CCL4, JUN, PLAUR and SCGN, increased less from baseline in SynF than in CtrlF infants at 2 months and 4 months (smaller change in SynF for all 16; nominal p < 0.05 for 13 of 16 at 4 months and 7 of 16 at 2 months, at both timepoints for CD6, CD70, PLAUR and SCGN; none significant after FDR correction) (**Fig. 5d**). The consistency of an attenuated-activation pattern across timepoints and among proteins with overlapping functions (cytotoxic/effector lymphocyte activation and inflammatory chemotaxis) indicate dampened inflammatory responses in SynF *vs* CtrlF feeding in this cohort, even when individual markers remain nominally significant only.

To test whether a coordinated biological process associated with feeding mode could be identified, we constructed a joint co-regulation network combining the 27 immune cell population frequencies (CLR-transformed) and all 358 Olink proteins (log_2_ NPX), using weighted gene co-regulation network analysis (WGCNA)^40^. This identified six co-regulation modules (**Fig. 5e**), numbered by size. Module 3 (40 features: CD8^+^ T central-memory, effector-memory, and TEMRA populations, together with 37 Olink proteins) captured a coherent CD8^+^ effector/memory T cell axis, including 7 of the 16 proteins highlighted above (KLRB1, CD160, TNFSF10, IL12RB1, FASLG, GZMA, LAMP3). Pairwise correlations among Module 3 features were substantially stronger than background (median bicor 0.43 *vs* 0.11 across the rest of the network), confirming these markers represent a coordinated biological unit during immune development. The Module 3 eigengene did not differ between feeding groups at baseline (Cohen’s d=0.25, p=0.368) or 2 months (d=0.05, p=0.879) but was significantly lower in SynF at 4 months (d=−0.44, p=0.027) (**Fig. 5f**). Collectively, these findings are consistent with SynF feeding promoting a modestly slower immune maturation trajectory, and more regulated immune activation both at the intestinal mucosa and in circulation.

### Vaccine antibody responses

To assess the impact of CtrlF *vs* SynF feeding on vaccine immunogenicity, we measured IgG antibodies against 36 vaccine-related antigens at 4 months. Vaccine-specific IgG responses did not differ meaningfully between feeding groups and none of the 36 antigens reached FDR significance (**Supplementary Fig. 5**). Four antigens showed nominally higher levels in CtrlF (p<0.05), but all antigens were from the Measles/Mumps/Rubella, MMR vaccine (measles nucleoprotein, rubella E1, native measles, mumps nucleoprotein) and these vaccines were not administered until the age of nine months. These antibodies therefore likely represent maternal antibodies transferred during pregnancy^41^, and higher levels at 4 months could reflect increased FcRn-mediated IgG recycling associated with systemic inflammation in CtrlF^42^.

### Trajectories of gut microbiome colonization track with intestinal inflammation

To go beyond individual microbe abundances and metrics of microbial diversity, we inferred Fecal Community Type (FCT) trajectories^43^. Four FCTs described trajectories (**Fig. 6a**) and FCTs were strongly associated with feeding group (**Fig. 6b**). CtrlF fed infants were predominantly assigned to trajectories T1 and T4, while SynF fed infants were predominantly assigned to trajectories T2 and T3, the latter characterized by early and sustained bifidobacterium dominance (data not shown). FCT trajectory was also associated with fecal markers of intestinal inflammation and barrier function across visits (**Fig. 6c**). Fecal pH differed most consistently and strongly between trajectories at every timepoint, secretory IgA showed trajectory-associated differences that shifted in magnitude and direction over time, and the antitrypsin-related measures showed more timepoint-specific differences, most prominent from 2 to 4 months. Together with the feeding-group associations in **Fig. 6a** and **b**, this supports gut microbial community trajectory as a route through which SynF feeding shapes the local intestinal environment and dampen inflammatory responses early in life.

**Figure 6.**
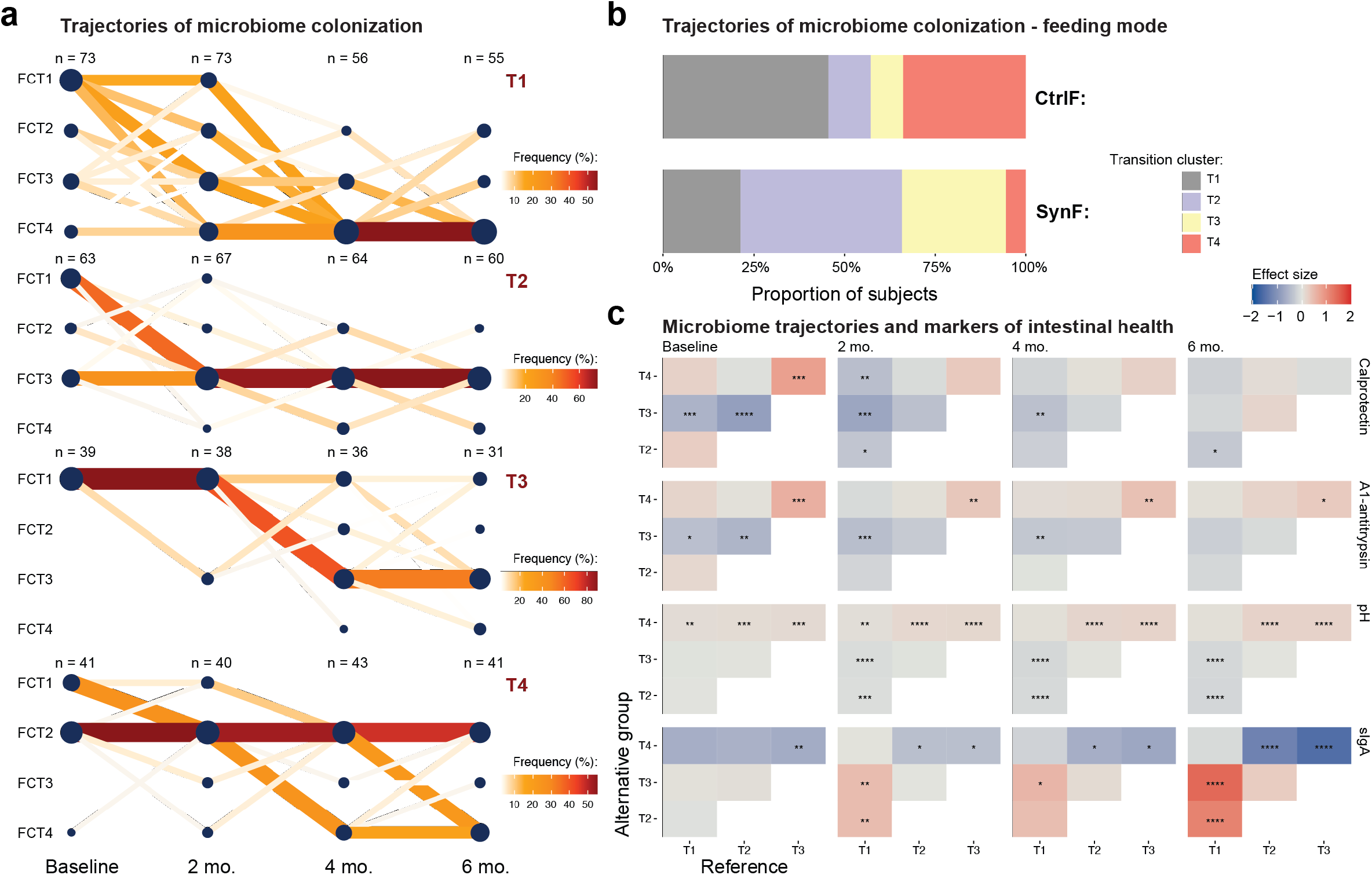
Pace of immune-microbe development in relation to feeding practices. a) Transition graphs of four clusters across fecal community types, FCTs and timepoints. The color and thickness of transition lines indicate number of samples associated with each trajectory, T1-4. b) Fraction of samples from children fed control formula (CtrlF) or Synbiotic formula (SynF), assigned to the four microbiome colonization trajectories in (a). c) associations between microbiome colonization trajectories and stool inflammatory biomarkers. On x-axis are reference trajectories and y-axis alternative trajectories and boxes colored by effect size differences and stars indicate adjusted p-values. * p<0.05, ** p<0.01, *** p<0.001, **** p<0.0001.

### Respiratory tract infections in relation to feeding groups

An increased propensity to respond with strong inflammatory responses upon infection has been linked to immune-microbiome interactions at mucosal surfaces, particularly in the airways^44–47^. To assess the effects of formula feeding mode and immune development on real-world consequences for health, we tracked episodes of upper respiratory infections (URTI) in infants fed CtrlF and SynF (**Fig. 7a-b**). We observed fewer episodes of URTI events in SynF *vs* CtrlF (Poisson regression, p=0.023) and lower Incidence Rate Ratios, IRR per child year in SynF *vs* CtrlF (IRR=0.586; 95% CI: 0.356, 0.954; p=0.023) corresponding to a 41% reduction in event rates (**Fig. 7c**). The duration of symptomatic infections is an indication of severity and linked to overall immune regulation. In this study, we found a trend towards shorter duration of first URTI in the SynF group compared to CtrlF (p=0.07)(**Fig. 7d**). These findings point towards a real-world consequence of a more tempered immune development and immune-microbe interactions less prone to drive inflammatory responses may be fewer symptomatic infections. Given that such infections have been linked to risks of immune mediated diseases such as asthma and wheeze, future trials with longer follow-up will be required to investigate whether synbiotic formula feeding and dampened inflammatory responses could prevent immune mediated disease.

**Figure 7.**
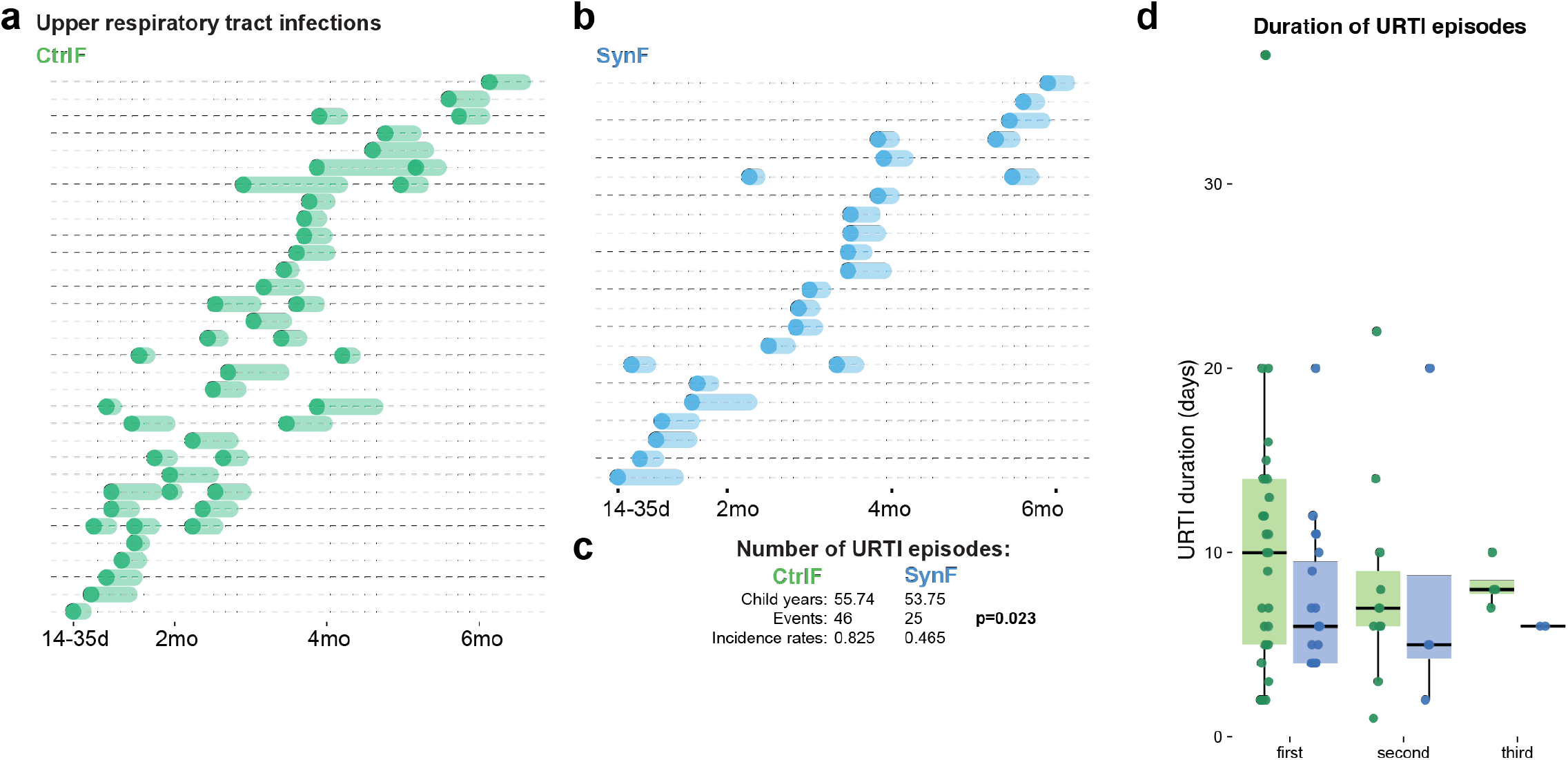
Upper respiratory tract infections in relation to feeding practices. a) Episodes in infants fed CtrlF. Each row represents a single child with reported upper respiratory infection (URTI) episodes. b) Same as (a) for infants fed experimental formula, SynF. c) Statistical comparison of the number of events (Poisson regression). d) Duration of reported episodes (days) by feeding group across first, second and third episodes.

## Discussion

This randomized, double-blind, controlled trial demonstrates that a synbiotic infant formula combining seven HMOs, BMOs, and *B. infantis* LMG11588 supports non-inferior growth, while promoting healthier microbiome and immune development. The prespecified primary and secondary outcomes for which the study was powered, converge on a coherent biological narrative, that synbiotic supplementation promotes a bifidobacterium-dominant gut microbiome which tempers inflammatory signaling both systemically and at mucosal sites, without compromising vaccine immunogenicity or infant health.

The primary and prespecified endpoint for this trial was growth (defined by weight gain velocity) non-inferiority, and our findings replicate the consistent safety signal seen in prior trials of the same probiotic strain^32^ and HMO supplementation trials^19–21^. What distinguishes this trial is the use of a synbiotic formulation and the deep analyses of microbiome composition, microbial metabolites, markers of intestinal health, and systems-level analyses of immune development in longitudinal blood samples. The results show robust *B. infantis* colonization in infants fed SynF as compared to CtrlF (approaching 100% *vs* <10%), exceeding what has been reported previously for probiotic or prebiotic interventions in formula-fed infants^48^. This also parallels our prior work in breastfed infants supplemented with *B. infantis,* which demonstrated that *B. infantis* colonization and HMO fermentation robustly correlated with increased tryptophan metabolites in general, and indole-3-lactic acid (ILA), in particular as a key immunomodulatory metabolite^8^, a finding replicated here in fully formula-fed infants. This result also suggests a mechanistic pathway for the observed beneficial effects whereby *Bifidobacterium*-dependent tryptophan fermentation leads to dampened inflammatory responses against colonizing microbes. In addition, the fecal markers of inflammation calprotectin and α-1-antitrypsin were lower whereas the secretory IgA produced by infants themselves in the absence of breastmilk was higher, indicating a less inflammatory gut environment and a healthier mucosal barrier in the gut.

The longitudinal effects on systemic immune development herein have not been reported previously in exclusively formula-fed infants. The observation that gut microbial community evolution tracked with feeding group, and with markers of intestinal inflammation, is mechanistically informative and consistent with a model in which immune- and barrier-related changes are mediated through effects on colonizing microbes rather than direct effects of formula components on immunity. Unsupervised network analysis (WGCNA) integrating the cell and protein data independently converged on a CD8⁺ T effector/memory axis as a co-regulated module (rather than a set of coincidental cell and protein hits) that was lower in SynF infants by 4 months after birth, lending additional support to a synbiotic-associated tempering of systemic immune activation and dampened inflammatory responses to commensal microbes. The finding of fewer URTI episodes and a trend towards a shorter duration of URTIs in the SynF group provides evidence that the immunological changes we observed may translate the optimized immune-microbiome interactions into a real-world health outcome of clinical relevance. This is also in line with data from a preclinical mouse model^49^. Mechanistically, it is tempting to speculate that rates of infection did not differ between groups but that inflammatory responses to colonizing microbes in the gut and perhaps also in the airways, leads to more balanced responses also upon viral infections and potentially also other environmental exposures such as food components, vaccines, and environmental chemicals, with less inflammation and tissue damage as a result. Future studies will be required to test this hypothesis directly and assess possible protection from immune mediated diseases such as asthma and allergy.

Several limitations deserve consideration. The trial was conducted exclusively in metropolitan Manila, the Philippines, in families who had independently elected not to breastfeed prior to enrollment. This appropriately restricts the trial to infants for whom formula feeding is the established nutritional mode but precludes an internal breastfed comparator arm. The degree of bridging achieved by the synbiotic formula can therefore only be inferred from the biomarker literature. The immune sub-study enrolled 86 of 240 infants on a first-come, first-served basis with possible selection bias. Mass cytometry and Olink findings are best considered exploratory as the large number of immune parameters evaluated means that independent replication is required for validation purposes. It is also important to understand how the same interventions replicate their effects across different ethnic populations and different environmental contexts, given the importance of microbial and other environmental exposures on immune variation in humans^50^. The URTI outcome was captured as an adverse event rather than a pre-specified primary endpoint, making it susceptible to differential ascertainment and recall bias across sites and visits. The trial was not powered to detect subgroup-specific effects, and therefore the stronger *Bifidobacterium* enrichment in C-section-born and sibling-free SynF fed infants, while biologically plausible, is exploratory. Additionally, the multi-component synbiotic design precludes disentangling the individual contributions of HMOs, bovine MOS, and *B. infantis,* a limitation that would require additional factorial or dose-finding trials to address, although several previous trials have been conducted on some of these components already. This trial demonstrates effects on growth, colonization, biomarkers, and exploratory immune measurements but does not establish any associations with long-term protection against allergy, infection, or immune-mediated disease and that remains to be tested?

The trial was conducted in a predominantly urban Philippine cohort with high rates of cesarean delivery and very low baseline *B. infantis* colonization among CtrlF fed infants (<10%). This baseline colonization ecology resembles the current situation in many high-income countries where generational depletion of *B. infantis* has occurred^51–53^, suggesting that the dramatic colonization effect is likely reproducible in such settings, but perhaps not in settings where baseline colonization is higher. However, differences in microbiome background, infection pressure, antibiotic exposure, and vaccine schedules across countries are likely to modulate the immunological effects observed, and findings cannot be assumed to generalize uniformly.

The restriction to term infants whose parents elected not to breastfeed means these findings do not extend to preterm or low birth weight populations, to mixed-fed infants, or to settings where breastfeeding is supplemented or discontinued for medical reasons. The 6-month observation window, while sufficient to characterize microbiome and immune development during the most dynamic period, does not allow conclusions about the durability of the immune phenotype after formula cessation or dietary diversification. Whether the dampened inflammatory trajectory translates into measurable reductions in atopic disease, autoimmunity, or metabolic outcomes in later childhood remains an important open question requiring extended follow-up.

In summary, this trial provides multi-modal evidence that a complex synbiotic formula can substantially recapitulate key features of the breastfed gut-immune axis in formula-fed term infants. The convergent evidence from microbiome sequencing, metabolomics, mucosal biomarkers, and systems-level immunomonitoring provides insights beyond what any single modality could establish. Future trials should prioritize replication in geographically diverse cohorts, pre-specified clinical endpoints for additional real-world health outcomes such as infections and atopic outcomes, and longer follow-up to determine whether early-life immune programming by synbiotic formula has lasting consequences for child health.

## Methods

### Study design and population

In this randomized, double-blind, controlled intervention study (ClinicalTrials.gov: NCT06073652), infants were recruited from 3 medical centers in the metropolitan Manila (Philippines) area between August 2023 and February 2024. Infants aged ≥14 to ≤35 days were enrolled if they met the following inclusion criteria: healthy term infant (≥37 weeks of gestation); birth weight ≥2500g and ≤4500g; and at the time of enrollment, infants were exclusively consuming and tolerating a cow’s milk infant formula and their parents have independently elected not to breastfeed before enrollment. Infants were excluded if they met any of the following criteria: evidence of feeding difficulties or formula intolerance such as vomiting or poor intake (investigator’s discretion); have conditions requiring infant feedings besides those specified in the protocol; have conditions or medical history that could interfere with the study conduct and interpretation such as major congenital malformations, suspected or documented systemic or congenital infections, or previous or ongoing severe medical or laboratory abnormality that could affect study participation; presently received or have received prior to enrollment medication(s) or supplement(s) which are known or suspected to affect fat digestion, absorption and/or metabolism, stool microbiota and characteristics, growth, or gastric acid secretion; and currently participating or have participated in another interventional clinical trial since birth. No changes were made to the trial methods, including design, eligibility criteria, or pre-specified outcomes after trial commencement.

### Ethics details

This study was approved by the Asian Hospital and Medical Center Research Ethics Committee (study 2022-038), the University of Perpetual Help System Institutional Ethics Review Board (study UPHS-IERB 2023-008), and the Ethics Review Committee of the Research Institute for Health Sciences at the University of the East Ramon Magsaysay Memorial Medical Center (study 1510/P/2023/082) and was conducted in compliance with International Conference on Harmonization Guidelines for Good Clinical Practice and the Declaration of Helsinki. Written informed consent was provided by the parent(s)/legally authorized representative(s) (LAR) prior to study enrollment.

### Randomization procedure and blinding

The infants who met the eligibility criteria were randomized to receive the synbiotic formula (SynF) or the control formula (CtrlF) in a 1:1 allocation ratio using the Medidata Randomization Trial Supply Management (New York, United States) with the dynamic allocation algorithm. The randomization was stratified by sex (female/male), center, and mode of delivery (vaginal/caesarean). Participants, parents/LAR(s), care providers at study sites, clinical investigators, laboratory personnel, data analysts, and sponsor personnel were all blinded to group assignments for the duration of the study. The randomization sequence was generated centrally by an independent biostatistician at Nestlé prior to study start and held within the Medidata RTSM. Study site investigators confirmed participant eligibility and formally enrolled each infant into the RTSM, which then automatically released the allocated formula code. No site personnel had access to the allocation sequence. The synbiotic and control formulas were identical in appearance, color, smell, packaging format, and labeling; labels indicated only the individualized product code, age range, and that the product was for clinical trial use, with no indication of group assignment.

### Intervention and study products

Infants began their intervention at V1 (baseline, 14-35 days after birth) and continued through V6 (6 months of postnatal age), with additional clinic visits at V2 (1.5 months), V3 (2 months), V4 (3 months), and V5 (4 months). The synbiotic formula (SynF) contained a standard bovine-milk based term infant formula with 13.4 g/L protein composed of 65% intact whey protein: 35% casein protein ratio, 100% lactose, and a fat blend containing 45% high palmitate in the sn-2 position with the remaining fat component composed of soy oil, sunflower oil and coconut oil. Additionally, the SynF was supplemented with a synbiotic blend which included 2.66 g/L of 7 manufactured HMOs (2’FL, 3FL, DFL, LNT, LNnT, 3’SL, 6’SL), 4.55 g/L bovine milk derived oligosaccharides (MOS; source of 3’GL, 4’GL, 6’GL), and 1×10^6 colony forming units (CFU) *B. infantis* (LMG11588) per gram formula powder. The control formula (CtrlF) group received the same formula as SynF except without the synbiotic blend (lactose was increased to compensate for the lack of HMOs and MOS). All study products were produced by Wyeth Nutritionals Ireland Ltd. (Askteaton, Co Limerick, Ireland) and provided in 400g cans and were fed orally, ad libitum, with amounts varying based on age, developmental stage, and appetite. Although the formula intervention ended at V6 (6 months of age), parents/LARs were invited to complete an infant illness questionnaire every 6 months until age 36 months. The trial was completed as planned. Recruitment ended after the pre-specified sample size had been enrolled; no, early stopping rules were triggered. This follow-up phase is still ongoing as of the date of finalization of this manuscript and these results will be published separately.

### Growth

Anthropometric measurements including weight (g), length (cm), and head circumference (cm) were collected using standardized procedures at V1 (baseline) through V6 (6 months). Weight gain measured as mean daily weight gain in g/day from V1 (baseline) to V5 (4 months) was the primary endpoint. Corresponding *z*-scores including weight-for-age, length-for-age, weight-for-length, and head-circumference-for-age were determined using the World Health Organization growth standards as the reference.

### Gastrointestinal tolerance

Gastrointestinal (GI) tolerance, symptoms, and related behaviors were examined using the GI Symptom and Behavior Diary and the Infant Gastrointestinal Symptom Questionnaire (IGSQ)^28^, a validated, standardized questionnaire with 13 items across 5 domains (crying, stooling, flatulence, fussiness, and vomiting/spitting up)^28^. Stool frequency and consistency and GI symptoms and related behaviors were recorded in the retrospective 1-day record at V1 (baseline) and prospective 3-day GI Symptom and Behavior Diary for 3 days prior to V2 (1.5 months), V3 (2 months), V4 (3 months), V5 (4 months), and V6 (6 months). Stool frequency (mean number of stools), stool consistency (mean score; 1=watery, 2=runny, 3=mushy/soft, 4=firm, 5=hard), GI symptoms including frequency and amount of spitting up/vomiting and flatulence, and GI-related behaviors such as crying, fussiness, and sleeping were captured in this diary. Additionally, the IGSQ was administered at V1 (baseline) and at V2 (1.5 months), V3 (2 months), V4 (3 months), V5 (4 months), and V6 (6 months) to derive a composite measure of GI symptom burden. A composite index score ranging from 13-65 was calculated by summing the domain scores, with lower scores representing a lower GI burden, so a score of 13-23 is considered indicative of good GI tolerance, a score of 24-30 indicates some GI distress, and scores above 30 are considered to reflect clinically meaningful GI symptom burden.

### Biological sample collection

Fecal samples were collected at home by the parent(s)/LAR(s) at V1 (baseline) after the actual visit but prior to start of product intake and on any of the three days prior to V3 (2 months), V5 (4 months), and V6 (6 months). The parent(s)/LAR(s) received collection kits and a manual instructing them how to collect, store, and transport the samples. Once the fecal samples were brought to the study site, they were kept frozen at −80°C until analysis. Blood samples were collected by a medical technologist using the appropriate equipment (e.g., microneedles) for infant blood sampling and/or according to the site’s standard of care. Samples of approximately 0.5-1.0 ml were collected at V1 (baseline), V3 (2 months), and V5 (4 months) in a subset of about 40 infants/randomized group on a first come, first serve principle so the first 40 parent(s)/LAR(s) in the SynF and CtrlF groups, respectively, to provide the optional consent for blood sample were included.

### Fecal microbiota analysis

Fecal microbiota composition was assessed using shotgun metagenomics as described^22^, with minor modifications. Briefly, deoxyribonucleic acid (DNA) was extracted from the fecal samples using the NucleoSpin Stool kit (Machery-Nagel) with bead beating at 2700 rpm for 5 min. DNA libraries were prepared using the Watchmaker DNA Library Prep Kit (7K0019-1K). Genomic DNA was fragmented using a mastermix of Watchmaker Frag/AT Buffer and Frag/AT Enzyme Mix. IDT xGen UDI Primers and IDT Stubby Adapters were added to each sample followed by 7 cycles of PCR to construct the DNA libraries. The final DNA libraries were purified using CleanNGS magnetic beads (CleanNA) and eluted in nuclease-free water. Following elution, the libraries were quantified using the Qubit™ fluorometer dsDNA HS Assay Kit and sequenced on an Illumina NovaSeq X platform at 2×150bp. After removal of adapter and human DNA, fecal microbiota composition and diversity metrics were determined using CHAMP^54^. Gut Metabolic Modules were annotated based on species-level KO annotations, assigning a GMM if it included at least 2/3 of the KOs needed to complete the functionality of the module^55^. For modules with multiple reaction paths, only one path was required to be 2/3 complete and for modules with less than four steps, all steps were required to be comprised in the given species. GMM abundance was calculated as the summed abundance of all species annotated with a given module. HMO-specific glycosyl hydrolase (GH) families were annotated using run_dbcan (v. 3.0.2)^56^, retaining annotations that were identified by both HMMER (E < 1e-15, coverage > 0.5) and DIAMOND (E < 1e-100). Each GH gene was classified as “secreted” if a signal peptide was detected by DeepSig (v. 1.2.5)^57^, or “internal” if no signal peptide was detected.

Microbiota diversity indices were calculated from rarefied species abundances using richness, Faith’s Phylogenetic Distance, and Shannon index as well as Bray-Curtis dissimilarity, and weighted UniFrac. Microbiome differences between the feeding groups were assessed using models correcting for site and baseline age using a linear regression framework with log-transformed relative abundances and a compositional bias correction based on LinDA^58^.

Fecal community types (FCT) were determined using Dirichlet Multinominal Mixture (DMM)^43^ at genus level with a prevalence filtering of >5% of samples and abundance filtering of >0.001% relative abundance. DMM models were run fitting from 1 to 10 clusters over 10 iterations using Laplace approximation to evaluate the optimal number of clusters. After an initial evaluation of five clusters that resulted in two being very similar, the model was constrained to four clusters. Individual subject trajectories of FCTs were established using state composition clustering. Other methods like Optimal Matching clustering were evaluated and dismissed due to the lack of a reference group. State Composition Clustering was done by summarizing each subject by the proportion of visits spent in each FCT state, computation of Manhattan distances between subjects on these proportions, and application of PAM (k-medoids) clustering. The optimal number of clusters was chosen based on Silhouette, Within-cluster sum of squares (WSS), and Gap statistics. Based on those three quality control measures, four trajectory clusters were identified as an optimal number.

### Fecal metabolic profile and markers of gut immune health and gut barrier function

Fecal pH was assessed using a pH meter (Orion, Thermo Scientific, USA). Fecal D- and L-lactate, D- and L-indole-lactate were measured by liquid chromatography coupled with a mass spectrometer (Acquity I-class UPLC Xevo TQ-XS system, Waters, USA). Briefly, fecal samples were weighed before and after lyophilization, and homogenized in orthophosphoric acid, followed by centrifugation to remove particulate matter. Supernatants were analyzed using LC–MS with separation of D/L-indole-3-lactic acid enantiomers using an Astec Chirobiotic R chiral column under a gentle gradient. Detection was carried out using a triple quadrupole mass spectrometer equipped with UniSpray^TM^ source technology operating in multiple reaction monitoring (MRM). Quantification of the 2 enantiomers was performed using internal calibration methodology^59^. Fecal short and branched chain fatty acids (propionate, butyrate, acetate, valerate, iso-butyrate, 2-methylbutyrate, iso-valerate) were measured by gas chromatography coupled with mass spectrometer (Agilent Technologies, USA) after deproteinization with 5-sulfosalicylic acid, chloroform extraction and derivatization with tert-butyldimethylsilylimidazole. Selectivity and detection were carried out on a single quadrupole mass spectrometer, followed by integration of chromatographic peaks with Openlab CDS3 (Agilent, USA) and data quantification on Slims (Agilent, USA).

Fecal secretory immunoglobulin A (sIgA, kit #K8870), calprotectin (kit #K6967, and α-1-antitrypsin (AAT, kit #K6750) were assessed by enzyme-linked immunosorbent assays (Immundiagnostik AG, Bensheim, Germany) according to manufacturers’ instructions.

Amounts of organic acids and protein markers were corrected per dry fecal weight and not per wet fecal weight as SynF had slightly softer stool.

### Blood sample processing

Blood was drawn into K_2_EDTA tubes and processed within 2 hours of collection. For immune cell analysis, 0.4 mL of whole blood was mixed 1:1 with a proprietary stabilizer (Cytodelics AB, Stockholm, Sweden)^60^ to preserve cell surface marker integrity. Plasma was separated by centrifugation and aliquoted (50 µL) for proteomic and antibody analyses. All samples were stored at −80°C and shipped on dry ice for centralized analysis.

### Whole blood analyses by mass cytometry (CyTOF)

Cryopreserved stabilized whole blood samples^60^ were thawed, fixed, and red blood cells were lysed according to the manufacturer’s protocol (Cytodelics AB). Cells were barcoded using the Cell-ID 20-Plex Barcoding Kit (Standard BioTools Inc., South San Francisco, CA) with an automated liquid handling system^61^ (Agilent Technologies) and pooled for batch staining. Cells were stained with a panel of >60 metal-conjugated antibodies targeting major immune cell lineages and functional subsets (**Supplementary Table 2**). Data were acquired on a CyTOF XT mass cytometer (Standard BioTools Inc.) at 300–500 cells/second using CyTOF software version 8.0. Raw data were normalized to calibration bead signals, arcsin h-transformed (co-factor 5), and batch-corrected using the ComBat algorithm (sva package in R). A manual gating strategy based on canonical marker combinations was used to define 28 core immune cell populations directly from the same normalized, batch-corrected data (**Supplementary Fig. 3**); this manually gated population set (27 populations after excluding one compositionally redundant population) was used for the primary cross-sectional and change-from-baseline population-frequency comparisons.

### Plasma proteomics (Olink Explore)

Plasma protein profiling was performed using the Olink Explore 384 Inflammation I Panel (Olink Proteomics, Uppsala, Sweden), which quantifies 384 inflammation-related proteins using Proximity Extension Assay (PEA) technology^39^ coupled with next-generation sequencing. The data were expressed as Normalized Protein Expression (NPX) values. After quality control filtering, 358 proteins were retained for analysis.

### Vaccine-specific antibodies (Luminex)

IgG antibodies against 36 vaccine-related antigens were measured using a custom suspension bead array on the Luminex FLEXMAP 3D platform. The panel included antigens from diphtheria, tetanus, pertussis (toxin, filamentous hemagglutinin, and pertactin), Haemophilus influenzae type b, hepatitis B, poliovirus (types 1–3), rotavirus, measles, mumps, and rubella vaccines (**Supplementary Table 3**). Bound IgG was detected using R-phycoerythrin-conjugated anti-human IgG, and median fluorescence intensity (MFI) values were z-score normalized for cross-sample comparison.

### Adverse events

Each principal investigator collected information pertaining to adverse events (AEs) or serious AEs (SAEs) on the electronic case record form (eCRF). Each AE was classified as mild, moderate, or severe and the relationship with the study product was noted as unrelated, unlikely, probable, or related. Additionally, medication type and duration of use including antibiotics and antipyretics were collected in concomitant medication reporting forms as part of the eCRF throughout the study period.

### Sample size

The sample size for this trial was determined using the primary endpoint of weight gain in g/day to 4 months of age and the key secondary endpoint of sIgA concentration per dry fecal weight at 4 months. To show a non-inferiority boundary of −3g/day with a power of 80%, 88 infants per arm would be required for the primary endpoint. Based on the differences in sIgA concentrations observed across groups in a previous study and an α-level of 5%, it was estimated that 108 infants per arm would be needed to demonstrate superiority of sIgA. With the assumption of a 10% attrition rate, each group would need 120 infants (240 in total). No pre-specified interim efficacy analyses were performed.

### Prespecified statistical analyses

Analyses were completed using several datasets. The intention-to-treat (ITT) analysis population consisted of all enrolled/randomized infants. The safety analysis set (SAF) comprised all infants in the ITT group who received at least one feeding of the study products. The full analysis set (FAS) included all infants from SAF except for those who could not satisfy study entry eligibility criteria or had no post-randomization data and were analyzed according to the assigned group per randomization. Lastly, the per-protocol (PP) population comprised of all FAS infants without any major departures from the protocol that could potentially influence the primary outcome. The primary endpoint of weight gain was analyzed in the FAS and PP populations while all secondary endpoints were evaluated in the FAS dataset. Safety analyses were conducted using the SAF population.

All variables were summarized using the appropriate descriptive statistics (e.g., mean, standard deviation [SD], frequency, percentage observed, etc.). The primary endpoint of weight gain was analyzed by an analysis of covariance (ANCOVA) model with adjustment for baseline weight, sex, and center. The key secondary endpoint of sIgA at V5 (4 months) was also analyzed using an ANCOVA model correcting for baseline value, sex, and center. Additional anthropometry measures (weight, length, head circumference, and corresponding *z*-scores), vital signs, GI tolerance, stool frequency and difficulty in passing stool, and stool consistency were examined using ANCOVA models at V2 (1.5 months), V3 (2 months), V4 (3 months), V5 (4 months), and V6 (6 months) with adjustment for baseline value, sex, and center. Metabolic profile and fecal markers of immune health and gut barrier function were analyzed at V3 (2 months), V5 (4 months), and V6 (6 months) while blood markers of systemic immunity were evaluated at V3 (2 months) and V5 (4 months). ANCOVA models correcting for baseline value, sex, and center were utilized for all three outcomes. Associations between microbiota and fecal biomarkers were done using Pearson correlations. Two-sided statistical tests with a significance level of 5% for nominal p-values and 10% for FDR (false discovery rate) correction were used across the different outcomes. SAS 9.4 (or higher) or R 4.1 (or higher) were used to perform statistical analyses.

### Blood immune marker analyses

For Mass cytometry and Olink data, cross-sectional comparisons between feeding groups at each time point were performed using Wilcoxon rank-sum tests. Longitudinal analyses were conducted using linear mixed-effects models with the formula: Outcome ∼ Feeding Group × Time Point + Mode of Delivery + (1|Subject), where Subject was included as a random intercept to account for repeated measures. Mode of delivery was included as a covariate given its known influence on early-life immune development. Multiple testing correction was applied using the Benjamini-Hochberg method; FDR-adjusted p<0.05 was considered statistically significant, while unadjusted p<0.05 was reported as exploratory. Effect sizes were calculated using Cohen’s d with 95% confidence intervals. For vaccine-specific antibodies, cross-sectional comparisons between feeding groups were performed at V5 (4 months) using Wilcoxon rank-sum tests. An integrated immune maturation metric (pseudotime) was derived by Slingshot trajectory analysis applied to the top 20% most time-varying cell populations and, separately, the top 20% most time-varying Olink plasma proteins, ordered by sampling age; resulting pseudotime values were z-scored and compared between feeding groups at each timepoint by Wilcoxon rank-sum and Mood’s median tests. To identify coordinated multi-parameter signatures, weighted gene co-regulation network analysis (WGCNA) was applied to a joint matrix combining the 27 manually gated CyTOF population frequencies (centered log-ratio, CLR, transformed) and all 358 Olink NPX values per sample (V1 (baseline), V3 (2 months), and V5 (4 months) pooled; n=248 samples with both modalities). A signed adjacency network was constructed using biweight midcorrelation (bicor) with a soft-thresholding power selected to approximate scale-free topology (R²≈0.8-0.9), followed by topological overlap matrix (TOM) transformation, average-linkage hierarchical clustering, and dynamic tree cutting (hybrid method, deepSplit=4, minimum module size=6) to define co-regulation modules; modules with highly correlated eigengenes (merge height <0.10) were combined, yielding six final modules numbered by decreasing size.

Each module’s eigengene (first principal component of its member features) was compared between feeding groups at each timepoint using the same Wilcoxon rank-sum test and Cohen’s d framework described above. Module membership (kME) — the correlation of each feature to its module’s eigengene — was used to rank features within each module (Fig. 5g).

## Acknowledgements

We sincerely thank all parents and caregivers who consented to their infants’ participation in the study. We highly appreciate the extended study teams at each site for their excellent contributions to study organization and conduct. We particularly thank Cecilia Fumero and Gemmie Low for assistance with study management, Niranjana Kodakuthumparambil and Danai Andreadi for assistance with data management and statistical analyses, Alric Mondragon for assistance with medical supervision, Ger Ryan and Elaine Kennedy for important contributions to study formula development and manufacture, and Gilles Le Carrou, Olivier Ciclet, Adrien Frezal for assistance with laboratory sample management, assay development, and lab analyses.

## Declaration

### Conflict of Interest

CKC, ND, HL, NS, NPH, and MN are employees of Société des Produits Nestlé S.A., the sponsors of this trial. JAR and JMM are employees of Cmbio, with whom Nestlé had a service agreement. The institutions of MRZC, MTA, and VMRTQ received grants from Nestlé to conduct the study. The investigational products were manufactured by Wyeth Nutrition (Askeaton, Ireland). Nestlé has a collaborative research agreement with Karolinska Institutet where the blood immune-related experimental analyses were conducted. PB and TL are cofounders of Cytodelics AB (Stockholm, Sweden) which produces and distributes whole blood cell stabilizer solutions used within this study. PB is a scientific advisor for Pixelgen Technologies AB, Helaina Inc., Scailyte AG, Oxford Immune Algorithmics Ltd., and the Swedish Olympic committee. PB is an executive board member of Sention Health AB, Stockholm, Sweden. The academic authors had full access to all the data in the study and had final responsibility for the decision to submit the manuscript for publication.

### Funding

This study was sponsored by Société des Produits Nestlé S.A. Scientists employed by Société des Produits Nestlé S.A. were involved in the study design, data collection, analyses, interpretation of the data, and writing and publication of the manuscript. The Brodin laboratory is supported by HORIZON-HLTH-2022-STAYHLTH-02 grant agreement 101094099 (INITIALISE), the Swedish research council (2019-01495, 2020-06190, 2020-02889, 2021-06529, 2021-05450, 2022-01567), The Swedish cancer society (CAN2015/587, CAN2018/764, 20 1175 PjF) and Knut & Alice Wallenberg Foundation (KAW2023-0344, VC-2021-0026, KAW 2020.0102), Göran Gustafsson Foundation (GG20200040), Swedish Society for Medical Research (SSMF, CG-22-0148-H-02) and Karolinska Institutet (2019-01019, 2018-02229). We thank SciLifeLab Affinity Proteomics facility Uppsala for Olink analyses.

### Ethics approval and consent to participate

This study was approved by the Asian Hospital and Medical Center Research Ethics Committee (study 2022-038), the University of Perpetual Help System Institutional Ethics Review Board (study UPHS-IERB 2023-008), and the Ethics Review Committee of the Research Institute for Health Sciences at the University of the East Ramon Magsaysay Memorial Medical Center (study 1510/P/2023/082) and conducted in compliance with the Declaration of Helsinki and the International Conference on Harmonization Guidelines for Good Clinical Practice. Parent(s)/legally authorized representative(s) provided written informed consent prior to study enrollment.

Trial registration: This trial was registered at ClinicalTrials.gov (NCT06073652).

### Protocol

The full study protocol and Statistical analysis plan are attached as supplementary material.

### Data availability

The individual participant data that underlie the results of this article, after de-identification, will be made available upon reasonable request to the corresponding author, subject to approval by Société des Produits Nestlé S.A. and applicable privacy regulations. De-identified processed data underlying the figures (immune cell frequencies, Olink NPX, vaccine IgG, anthropometrics, GI tolerance, adverse events, fecal biomarkers, metabolites and microbiome tables) are provided as Supplementary Data with blinded subject identifiers. The processed data underlying the immune analyses (Figs. 2 and 5 and Suppl. Figs. 4–5) are also available, together with the analysis code, at https://github.com/Brodinlab/barcenilla-et-al-synbiotic-formula-rct. Raw mass cytometry (FCS) and Olink data are not publicly deposited and are available upon reasonable request. **Open science**: Statistical analysis code and data is available on GitHub: https://github.com/Brodinlab/barcenilla-et-al-synbiotic-formula-rct. The Statistical analysis plan was pre-specified prior to database lock and is shared as supplementary material.

Participants and public involvement: Patients and/or the public were not involved in the design, conduct, reporting, or dissemination plans of this research.

## SUPPLEMENTAL MATERIALS

**Supplementary Fig. 1. Bifidobacterium colonization by mode of delivery and presence of siblings.** Relative abundance of Bifidobacterium (%, log scale) in stool of CtrlF (green) and SynF (blue) infants in the full analysis set at baseline, 2, 4 and 6 months of age. a) Infants born by C-section (top) or vaginal delivery (bottom). b) Infants with (top) or without (bottom) older siblings. Boxes show the median and interquartile range, and points are individual infants. P-values compare SynF with CtrlF at each timepoint.

**Supplementary Fig. 2. Stool propionate and its association with the gut microbiome.** Stool propionate measured at the same timepoints as the metagenomic sequencing by targeted mass spectrometry. The heatmap shows the correlation coefficient between each bacterial species and propionate at 0, 2, 4 and 6 months (dots mark significant correlations); the adjacent panel shows the group with the highest value for each species (CtrlF, green; SynF, blue; grey, non-significant association; white, no statistical result). Acetate and butyrate are shown in Fig. 4e.

**Supplementary Fig. 3. Mass cytometry gating strategy.** Manual gating hierarchy used to classify immune cell populations in whole blood, shown for one representative sample. Numbers are the percentage of events within the parent gate. CD45+ leukocytes were divided into neutrophils and non-neutrophils; eosinophils, B cells and T cells were then identified. The innate/myeloid branch resolves NK-cell subsets (CD56bright CD16−, CD56dim CD16+), classical, intermediate and non-classical monocytes, mDCs, basophils and pDCs. The T-cell branch resolves CD4+ and CD8+ T cells, HLA-DR+ T cells and TCRγδ T cells, with naive, central memory (TCM), effector memory (TEM) and TEMRA subsets defined by CD27 and CD45RA.

**Supplementary Fig. 4. Manually gated CyTOF cell population frequencies over time, by feeding group.** One panel per manually gated immune cell population (28 populations, % of CD45+ cells, plate-corrected), showing individual-infant trajectories (thin lines) at baseline, 2 and 4 months, colored by feeding group (CtrlF, green; SynF, blue). Bold lines with points show group medians. One compositionally redundant population was excluded from the analyses in Fig. 5; all 28 are shown here.

**Supplementary Fig. 5. Vaccine-specific IgG at 4 months.** Difference in IgG between SynF and CtrlF infants at 4 months for 36 vaccine-related antigens: Cohen’s d (x-axis, positive = higher in SynF) against nominal significance (−log10 p-value, y-axis). The dashed horizontal line marks nominal p = 0.05 and the dashed vertical line d = 0. Antigens are colored by category: infant-schedule vaccine (blue), maternal/MMR-family (pink) and non-vaccine pathogen (grey). Selected antigens are labelled. None of the 36 antigens reached significance after FDR correction.

**Supplementary Table 1.**
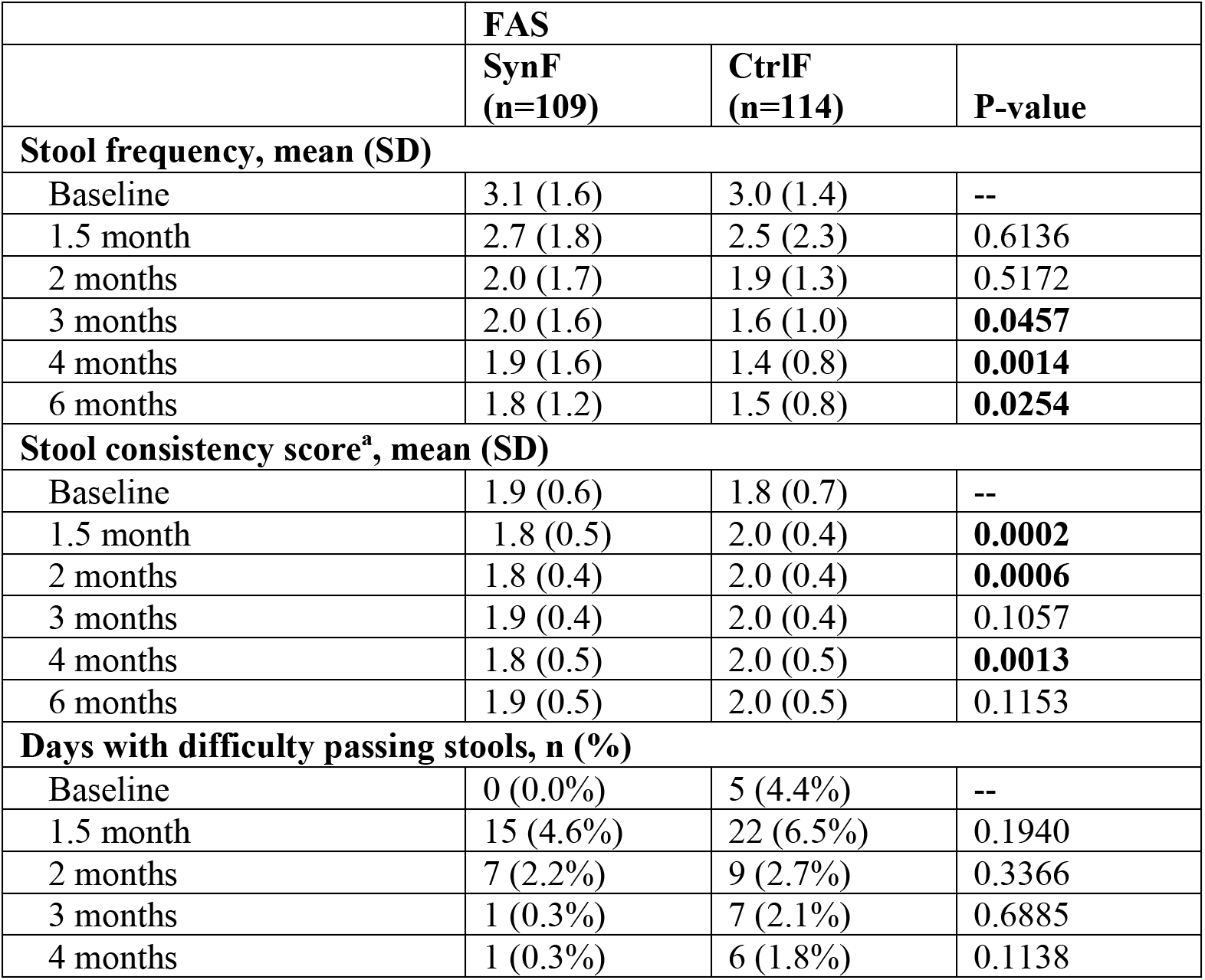
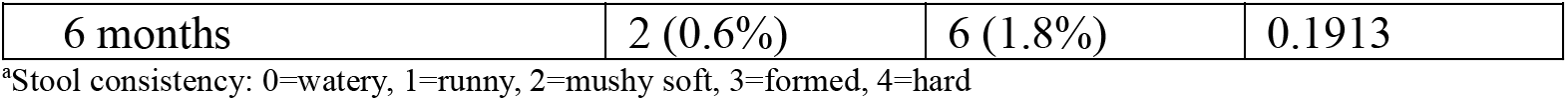
Stool characteristics.

**Supplementary Table 2.** Mass cytometry antibody panel.

| Metal | Marker | Clone | Source |
|---|---|---|---|
| 89Y | CD45 | HI30 | Standard BioTools |
| 110Cd | CD33 | WM53 | BioLegend |
| 111Cd | CD26 | BA5b | BioLegend |
| 112Cd | CD11c | Bu15 | BioLegend |
| 113Cd | IgD | IA6-2 | BioLegend |
| 114Cd | HLA-DR | L243 | BioLegend |
| 115In | CD57 | HCD57 | BioLegend |
| 140Ce | CD71 | CY1G4 | BioLegend |
| 141Pr | CD49d | 9F10 | Standard BioTools |
| 142Nd | CD43 | 84-3C1 | eBiosciences |
| 143Nd | CD3e | UCHT1 | BioLegend |
| 144Nd | CD15 | W6D3 | BioLegend |
| 145Nd | CD81 | 5A6 | BioLegend |
| 146Nd | CD52 | HI186 | BioLegend |
| 147Sm | CD1c | L161 | BioLegend |
| 148Nd | CD55 | JS11 | BioLegend |
| 149Sm | CD25 | 2A3 | Standard BioTools |
| 150Nd | CD64 | 10.1 | BioLegend |
| 151Eu | CD123 | 6H6 | BioLegend |
| 152Sm | TCRγδ | 5A6.E9 | Fisher Scientific |
| 153Eu | Siglec-8 | 837535 | R&D Systems |
| 154Sm | CD95 | DX2 | BioLegend |
| 155Gd | CX3CR1 | 8E10.D9 | BioLegend |
| 156Gd | CD20 | 2H7 | BioLegend |
| 157Gd | CD9 | SN4 C3-3A2 | eBiosciences |
| 158Gd | KIR (pooled) | Multiple | Multiple |
| 159Tb | CD22 | HIB22 | BioLegend |
| 160Gd | CD14 | M5E2 | BioLegend |
| 161Dy | CD161 | HP-3G10 | BioLegend |
| 162Dy | CD29 | TS2/16 | BioLegend |
| 163Dy | 4-1BB (CD137) | 4B4-1 | BioLegend |
| 164Dy | CD10 | HI10a | BioLegend |
| 165Ho | CD127 | A019D5 | Standard BioTools |
| 166Er | CD24 | ML5 | BioLegend |
| 167Er | CD27 | L128 | BioLegend |
| 168Er | CD141 | M80 | BioLegend |
| 169Tm | CD45RA | HI100 | Standard BioTools |
| 170Er | CD38 | HIT2 | BioLegend |
| 171Yb | CD85j | GHI/75 | BioLegend |
| 172Yb | CD147 | HIM6 | BioLegend |
| 173Yb | CD56 | NCAM16.2 | BD Biosciences |
| 174Yb | CD99 | HCD99 | BioLegend |
| 175Lu | CD28 | CD28.2 | BioLegend |
| 176Yb | CD39 | A1 | BioLegend |
| 191Ir | DNA Intercalator | — | Standard BioTools |
| 193Ir | DNA Intercalator | — | Standard BioTools |
| 194Pt | CD8a | SK1 | BD Biosciences |
| 195Pt | CD5 | UCHT2 | BioLegend |
| 196Pt | CD7 | CD7-6B7 | BioLegend |
| 198Pt | CD4 | RPA-T4 | BioLegend |
| 209Bi | CD16 | 3G8 | Standard BioTools |
| 102-108Pd | Barcoding | — | Standard BioTools |

*Note: KIR markers (158Gd) include pooled antibodies against KIR2DL2/L3, KIR2DL1/KIR2DS5, KIR3DL2, KIR3DL1, and KIR2DL5.*

**Supplementary Table 3.** Vaccine antigen panel.

| **Bead ID** | **Antigen** | **Vaccine/Pathogen** | **Coupling Method** |
|---|---|---|---|
| 193 | Measles Virus Nucleoprotein (HEK293) | Measles | NHS/EDC |
| 194 | Rotavirus VP7 Protein | Rotavirus | NHS/EDC |
| 199 | Rotavirus (Strain SA-11) | Rotavirus | NHS/EDC |
| 200 | Native Measles virus | Measles | NHS/EDC |
| 204 | Mumps Virus Nucleoprotein Recombinant | Mumps | NHS/EDC |
| 205 | Cytomegalovirus glycoprotein B (gB) | CMV | NHS/EDC |
| 207 | RSV A Glycoprotein G | RSV | NHS/EDC |
| 208 | B. pertussis toxin (mutant) | Pertussis | NHS/EDC |
| 209 | Tetanus Toxoid | Tetanus | NHS/EDC |
| 210 | HPV type 16 L1 Protein | HPV | NHS/EDC |
| 211 | HPV type 18 L1 Protein | HPV | NHS/EDC |
| 216 | Rubella E1 | Rubella | NHS/EDC |
| 217 | Rubella virus E1, C-terminal SHFc-tag | Rubella | NHS/EDC |
| 218 | Tetanus Toxoid, Recombinant Heavy Chain Fragment C | Tetanus | NHS/EDC |
| 219 | Rubella Spike Ectodomain (E1-E2) | Rubella | NHS/EDC |
| 224 | Mumps virus nucleoprotein, inactivated | Mumps | NHS/EDC |
| 225 | Native Mumps virus | Mumps | NHS/EDC |
| 231 | HBV Surface Antigen (subtype adw) | Hepatitis B | NHS/EDC |
| 232 | B. pertussis whole-cell (strain Tahoma I) | Pertussis | NHS/EDC |
| 233 | HPV type 33 L1 protein (VLP) | HPV | NHS/EDC |
| 234 | Poliovirus type 2 VP3-VP1 capsid | Polio | NHS/EDC |
| 235 | Poliovirus type 3 VP3-VP1 capsid | Polio | NHS/EDC |
| 236 | Epstein Barr Virus gp125 | EBV | NHS/EDC |
| 237 | S. pneumoniae Cell Wall Polysaccharide | Pneumococcus | NHS/EDC |
| 244 | B. pertussis Filamentous Hemagglutinin (FHA) | Pertussis | Mix&Go |
| 248 | Rubella virus nucleoprotein, C-terminal His-tag | Rubella | Mix&Go |
| 249 | Rubella Virus Grade 4, natural antigen | Rubella | Mix&Go |
| 250 | B. pertussis FHA - Bulk antigen | Pertussis | Mix&Go |
| 256 | Diphtheria mutated toxin | Diphtheria | Mix&Go |
| 257 | Diphtheria Toxoid | Diphtheria | Mix&Go |
| 259 | B. pertussis Pertactin Protein | Pertussis | Mix&Go |
| 264 | Mumps virus nucleoprotein | Mumps | Mix&Go |
| 265 | B. pertussis FHA - Native antigen | Pertussis | Mix&Go |
| 266 | B. pertussis whole-cell (strain Tahoma I) | Pertussis | Mix&Go |
| 267 | HPV type 6 L1 protein (VLP) | HPV | Mix&Go |
| 274 | Poliovirus type 1 Capsid protein (strain Sabin) | Polio | Mix&Go |

*Note: Control beads (bare beads, goat IgG, mouse IgG, and anti-human IgG) were included for each coupling method.*

## Notes

### Competing Interest Statement

CKC, ND, HL, NS, NPH, and MN are employees of Societe des Produits Nestle S.A., the sponsors of this trial. JAR and JMM are employees of Cmbio, with whom Nestle had a service agreement. The institutions of MRZC, MTA, and VMRTQ received grants from Nestle to conduct the study. The investigational products were manufactured by Wyeth Nutrition (Askeaton, Ireland). Nestle has a collaborative research agreement with Karolinska Institutet where the blood immune-related experimental analyses were conducted. PB and TL are cofounders of Cytodelics AB (Stockholm, Sweden) which produces and distributes whole blood cell stabilizer solutions used within this study. PB is a scientific advisor for Pixelgen Technologies AB, Helaina Inc., Scailyte AG, Oxford Immune Algorithmics Ltd., and the Swedish Olympic committee. PB is an executive board member of Sention Health AB, Stockholm, Sweden. The academic authors had full access to all the data in the study and had final responsibility for the decision to submit the manuscript for publication.

### Clinical Trial

NCT06073652

### Summary of Updates

Changes to analyses and language describing the conclusions. Author changes that better reflect the updated manuscript.

## REFERENCES

1. Victora, C. G. et al. Breastfeeding in the 21st century: epidemiology, mechanisms, and lifelong effect. Lancet 387, 475–490 (2016).

2. Manzoni, P. et al. Role of Lactoferrin in Neonates and Infants: An Update. Am. J. Perinatol. 35, 561–565 (2018).

3. Verhasselt, V., Tellier, J., Carsetti, R. & Tepekule, B. Antibodies in breast milk: Pro-bodies designed for healthy newborn development. Immunol. Rev. 328, 192–204 (2024).

4. Ballard, O. & Morrow, A. L. Human Milk Composition Nutrients and Bioactive Factors. Pedi-atr. Clin. North Am. 60, 49–74 (2013).

5. Bode, L. Human milk oligosaccharides: Every baby needs a sugar mama. Glycobiology 22, 1147–1162 (2012).

6. Sela, D. A. et al. The genome sequence of Bifidobacterium longum subsp. infantis reveals ad-aptations for milk utilization within the infant microbiome. Proc National Acad Sci 105, 18964–18969 (2008).

7. Underwood, M. A., German, J. B., Lebrilla, C. B. & Mills, D. A. Bifidobacterium longum subspecies infantis: champion colonizer of the infant gut. Pediatr Res 77, 229–235 (2015).

8. Henrick, B. M. et al. Bifidobacteria-mediated immune system imprinting early in life. Cell 10.1016/j.cell.2021.05.030 (2021) doi:10.1016/j.cell.2021.05.030.

9. Arrieta, M.-C. et al. Early infancy microbial and metabolic alterations affect risk of childhood asthma. Science Translational Medicine 7, 307ra152–307ra152 (2015).

10. Ho, N. T. et al. Meta-analysis of effects of exclusive breastfeeding on infant gut microbiota across populations. Nat. Commun. 9, 4169 (2018).

11. Bäckhed, F. et al. Dynamics and Stabilization of the Human Gut Microbiome during the First Year of Life. Cell Host Microbe 17, 690–703 (2015).

12. Goehring, K. C. et al. Similar to Those Who Are Breastfed, Infants Fed a Formula Contain-ing 2′-Fucosyllactose Have Lower Inflammatory Cytokines in a Randomized Controlled Trial 1 2 3 4. J. Nutr. 146, 2559–2566 (2016).

13. Forbes, J. D. et al. Association of Exposure to Formula in the Hospital and Subsequent Infant Feeding Practices With Gut Microbiota and Risk of Overweight in the First Year of Life. JAMA Pediatr. 172, e181161–e181161 (2018).

14. Thurl, S., Munzert, M., Boehm, G., Matthews, C. & Stahl, B. Systematic review of the con-centrations of oligosaccharides in human milk. Nutr. Rev. 75, 920–933 (2017).

15. Garrido, D., Kim, J. H., German, J. B., Raybould, H. E. & Mills, D. A. Oligosaccharide Binding Proteins from Bifidobacterium longum subsp. infantis Reveal a Preference for Host Gly-cans. PLoS ONE 6, e17315 (2011).

16. Bode, L. The functional biology of human milk oligosaccharides. Early Hum. Dev. 91, 619–622 (2015).

17. Donovan, S. M. & Comstock, S. S. Human Milk Oligosaccharides Influence Neonatal Muco-sal and Systemic Immunity. Ann. Nutr. Metab. 69, 41–51 (2017).

18. Sprenger, N., Tytgat, H. L. P., Binia, A., Austin, S. & Singhal, A. Biology of human milk oli-gosaccharides: From basic science to clinical evidence. J. Hum. Nutr. Diet. 35, 280–299 (2022).

19. Puccio, G. et al. Effects of Infant Formula With Human Milk Oligosaccharides on Growth and Morbidity. J. Pediatr. Gastroenterol. Nutr. 64, 624–631 (2017).

20. Marriage, B. J., Buck, R. H., Goehring, K. C., Oliver, J. S. & Williams, J. A. Infants Fed a Lower Calorie Formula With 2′FL Show Growth and 2′FL Uptake Like Breast-Fed Infants. J. Pediatr. Gastroenterol. Nutr. 61, 649–658 (2015).

21. Bosheva, M. et al. Infant Formula With a Specific Blend of Five Human Milk Oligosaccha-rides Drives the Gut Microbiota Development and Improves Gut Maturation Markers: A Ran-domized Controlled Trial. Front. Nutr. 9, 920362 (2022).

22. Picaud, J.-C. et al. Partially hydrolyzed, whey-based infant formula with six human milk oli-gosaccharides, B. infantis LMG11588, and B. lactis CNCM I-3446 is safe, well tolerated, and improves gut health: a staged analysis of a randomized trial. Front. Nutr. 12, 1628847 (2025).

23. Gopal, P. K. & Gill, H. S. Oligosaccharides and glycoconjugates in bovine milk and colos-trum. Br. J. Nutr. 84, 69–74 (2000).

24. Estorninos, E. et al. Infant formula containing bovine milk-derived oligosaccharides supports age-appropriate growth and improves stooling pattern. Pediatr. Res. 91, 1485–1492 (2022).

25. Laursen, M. F. et al. Bifidobacterium species associated with breastfeeding produce aromatic lactic acids in the infant gut. Nat Microbiol 6, 1367–1382 (2021).

26. Frese, S. A. et al. Persistence of Supplemented Bifidobacterium longum subsp. infantis EVC001 in Breastfed Infants. Msphere 2, e00501–17 (2017).

27. Capeding, M. R. Z. et al. Safety, efficacy, and impact on gut microbial ecology of a Bifidobacterium longum subspecies infantis LMG11588 supplementation in healthy term in-fants: a randomized, double-blind, controlled trial in the Philippines. Front. Nutr. 10, 1319873 (2023).

28. Riley, A. W., Trabulsi, J., Yao, M., Bevans, K. B. & DeRusso, P. A. Validation of a Parent Report Questionnaire. Clin. Pediatr. 54, 1167–1174 (2015).

29. Donald, K., Petersen, C., Turvey, S. E., Finlay, B. B. & Azad, M. B. Secretory IgA: Linking microbes, maternal health, and infant health through human milk. Cell Host Microbe 30, 650–659 (2022).

30. Gopalakrishna, K. P. et al. Maternal IgA protects against the development of necrotizing en-terocolitis in preterm infants. Nat. Med. 25, 1110–1115 (2019).

31. Ogawa, K., Ben, R. A., Pons, S., Paolo, M. I. L. & Fernández, L. B. Volatile Fatty Acids, Lactic Acid, and pH in the Stools of Breast-Fed and Bottle-Fed Infants. J. Pediatr. Gastroen-terol. Nutr. 15, 248–252 (1992).

32. Henrick, B. M. et al. Elevated Fecal pH Indicates a Profound Change in the Breastfed Infant Gut Microbiome Due to Reduction of Bifidobacterium over the Past Century. Msphere 3, e00041–18 (2018).

33. Brodin, P. Immune-microbe interactions early in life: A determinant of health and disease long term. Science 376, 945–950 (2022).

34. Fukuda, S. et al. Bifidobacteria can protect from enteropathogenic infection through produc-tion of acetate. Nature 469, 543–547 (2011).

35. Roduit, C. et al. High levels of butyrate and propionate in early life are associated with pro-tection against atopy. Allergy 74, 799–809 (2019).

36. Shenhav, L. et al. Microbial colonization programs are structured by breastfeeding and guide healthy respiratory development. Cell 187, 5431–5452.e20 (2024).

37. Olin, A. et al. Stereotypic Immune System Development in Newborn Children. Cell 174, 1277–1292.e14 (2018).

38. Street, K. et al. Slingshot: cell lineage and pseudotime inference for single-cell transcriptom-ics. BMC Genom. 19, 477 (2018).

39. Fredriksson, S. et al. Protein detection using proximity-dependent DNA ligation assays. Na-ture Biotechnology 20, 473–477 (2002).

40. Zhang, B. & Horvath, S. A General Framework for Weighted Gene Co-Expression Network Analysis. Stat. Appl. Genet. Mol. Biol. 4, Article17 (2005).

41. Pou, C. et al. The repertoire of maternal anti-viral antibodies in human newborns. Nature Medicine 1–6 (2019) doi:10.1038/s41591-019-0392-8.

42. Liu, X. et al. NF-κB Signaling Regulates Functional Expression of the MHC Class I-Related Neonatal Fc Receptor for IgG via Intronic Binding Sequences. J. Immunol. 179, 2999–3011 (2007).

43. Holmes, I., Harris, K. & Quince, C. Dirichlet Multinomial Mixtures: Generative Models for Microbial Metagenomics. PLoS ONE 7, e30126 (2012).

44. Pattaroni, C. et al. Bacterial communities co-develop with respiratory immunity early in life, linking dysbiosis to systemic monocyte signature and wheezing. Sci. Adv. 11, eadw1410 (2025).

45. Gutierrez, M. W., Bernardes, E. van T., Changirwa, D., McDonald, B. & Arrieta, M.-C. “Molding” immunity—modulation of mucosal and systemic immunity by the intestinal mycobi-ome in health and disease. Mucosal Immunol. 15, 573–583 (2022).

46. Kristensen, M. et al. The respiratory microbiome is linked to the severity of RSV infections and the persistence of symptoms in children. Cell Rep. Med. 5, 101836 (2024).

47. Indrio, F. et al. Health Effects of Infant Formula Supplemented with Probiotics or Synbiotics in Infants and Toddlers: Systematic Review with Network Meta-Analysis. Nutrients 14, 5175 (2022).

48. Closa-Monasterolo, R. et al. Safety and efficacy of inulin and oligofructose supplementation in infant formula: Results from a randomized clinical trial. Clin. Nutr. 32, 918–927 (2013).

49. Rashid, R. B. et al. Early-life immune imprinting by a B. infantis-HMO synbiotic shapes in-fant immune signatures and protects against experimental respiratory disease. Cell Rep. Med. 7, 102935 (2026).

50. Brodin, P. et al. Variation in the Human Immune System Is Largely Driven by Non-Heritable Influences. Cell 160, 37–47 (2015).

51. Olm, M. R. et al. Robust variation in infant gut microbiome assembly across a spectrum of lifestyles. Science 376, 1220–1223 (2022).

52. Jarman, J. B. et al. Bifidobacterium deficit in United States infants drives prevalent gut dysbiosis. Commun. Biol. 8, 867 (2025).

53. Taft, D. H. et al. Bifidobacterium Species Colonization in Infancy: A Global Cross-Sectional Comparison by Population History of Breastfeeding. Nutrients 14, 1423 (2022).

54. Pita, S. et al. CHAMP delivers accurate taxonomic profiles of the prokaryotes, eukaryotes, and bacteriophages in the human microbiome. Front. Microbiol. 15, 1425489 (2024).

55. Vieira-Silva, S. et al. Species–function relationships shape ecological properties of the hu-man gut microbiome. Nat. Microbiol. 1, 16088 (2016).

56. Zhang, H. et al. dbCAN2: a meta server for automated carbohydrate-active enzyme annota-tion. Nucleic Acids Res. 46, W95–W101 (2018).

57. Savojardo, C., Martelli, P. L., Fariselli, P. & Casadio, R. DeepSig: deep learning improves signal peptide detection in proteins. Bioinformatics 34, 1690–1696 (2017).

58. Zhou, H., He, K., Chen, J. & Zhang, X. LinDA: linear models for differential abundance analysis of microbiome compositional data. Genome Biol. 23, 95 (2022).

59. Visconti, G., Boccard, J., Feinberg, M. & Rudaz, S. From fundamentals in calibration to modern methodologies: A tutorial for small molecules quantification in liquid chromatography–mass spectrometry bioanalysis. Anal. Chim. Acta 1240, 340711 (2023).

60. Brodin, P., Duffy, D. & Quintana-Murci, L. A Call for Blood—In Human Immunology. Im-munity 50, 1335–1336 (2019).

61. Mikes, J., Olin, A., Lakshmikanth, T., Chen, Y. & Brodin, P. Mass Cytometry, Methods and Protocols. Methods Mol Biology Clifton N J 1989, 111–123 (2019).

