## Supplemental Fig 1 for "A Synbiotic Infant Formula Promotes Balanced Immune-Microbiome Interactions in Term Infants: A Randomized Controlled Trial"

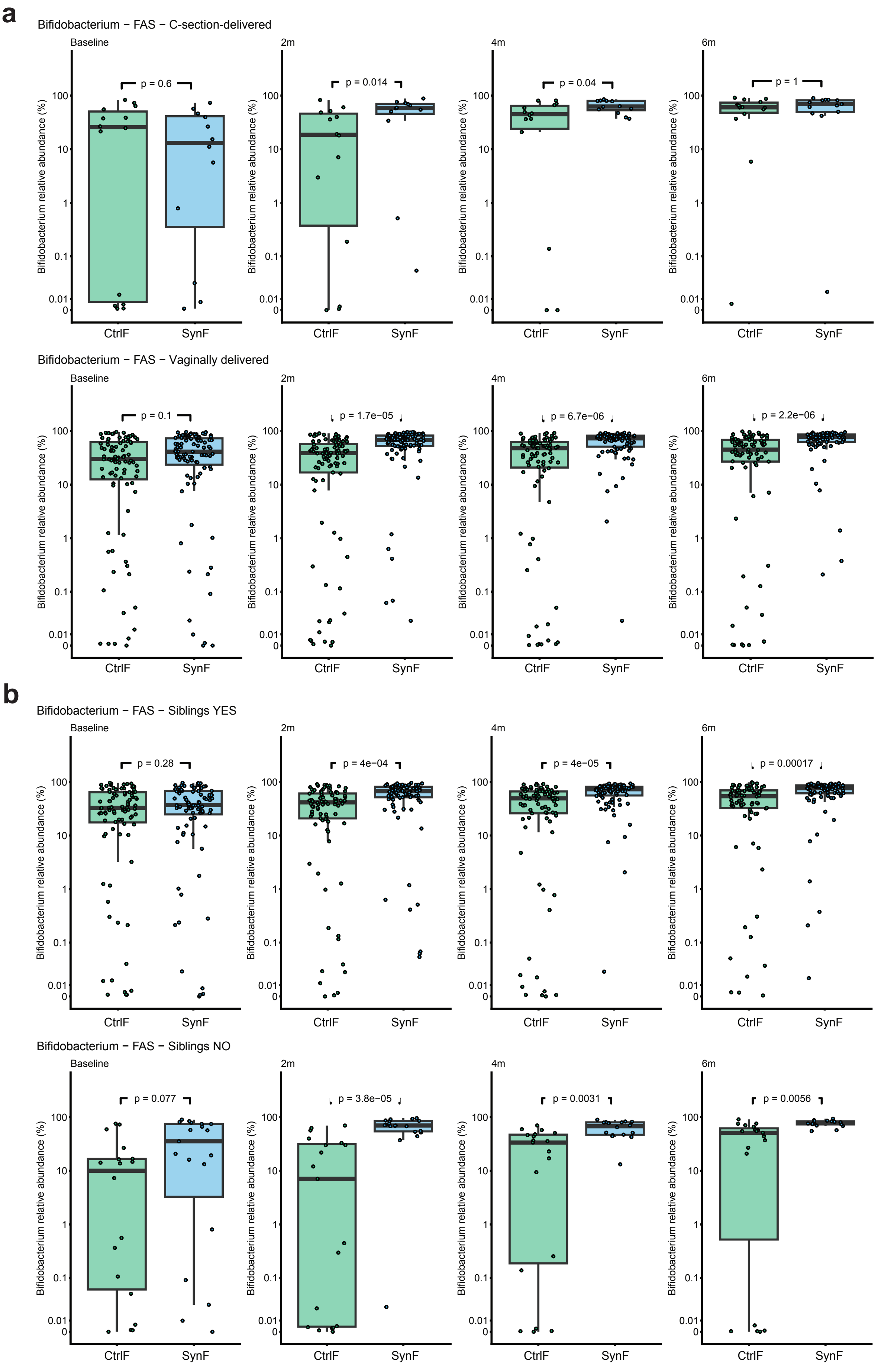


**Supplementary Fig 1. Bifidobacterium colonization a**) Relative abundances of Bifidobacterium at baseline, 2-, 4- and 6 months of age in stool from children born by C-section or Vaginal delivery. **b**) Same as in (a) but divided by siblings (yes or no).
