## Supplemental Fig 2 for "A Synbiotic Infant Formula Promotes Balanced Immune-Microbiome Interactions in Term Infants: A Randomized Controlled Trial"

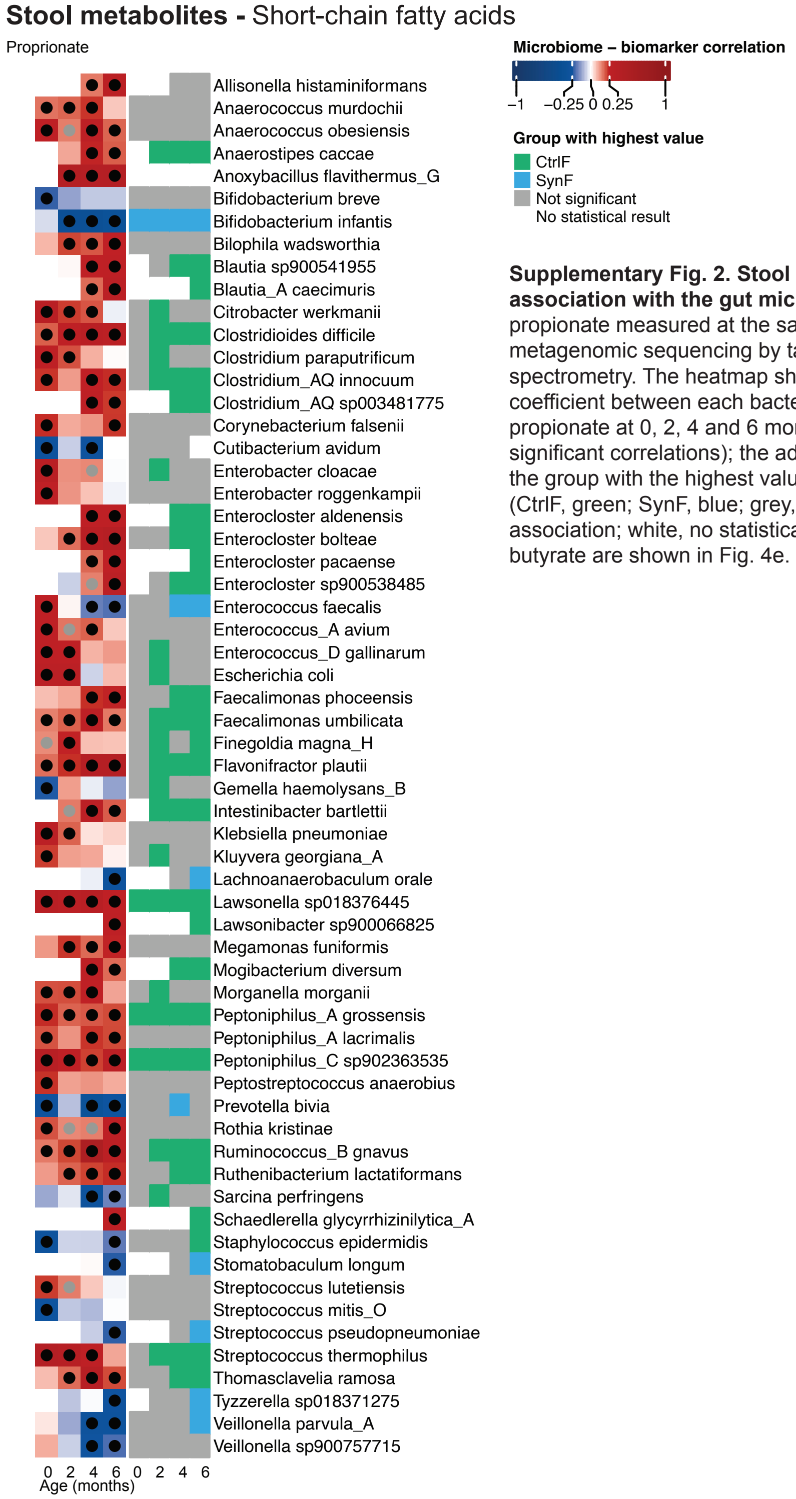


**Supplementary Fig. 2. Stool propionate and its association with the gut microbiome.** Stool propionate measured at the same timepoints as the metagenomic sequencing by targeted mass spectrometry. The heatmap shows the correlation coefficient between each bacterial species and propionate at 0, 2, 4 and 6 months (dots mark significant correlations); the adjacent panel shows the group with the highest value for each species (CtrlF, green; SynF, blue; grey, non-significant association; white, no statistical result). Acetate and butyrate are shown in Fig. 4e.
