## Supplemental Fig 3 for "A Synbiotic Infant Formula Promotes Balanced Immune-Microbiome Interactions in Term Infants: A Randomized Controlled Trial"

### Mass cytometry gating strategy

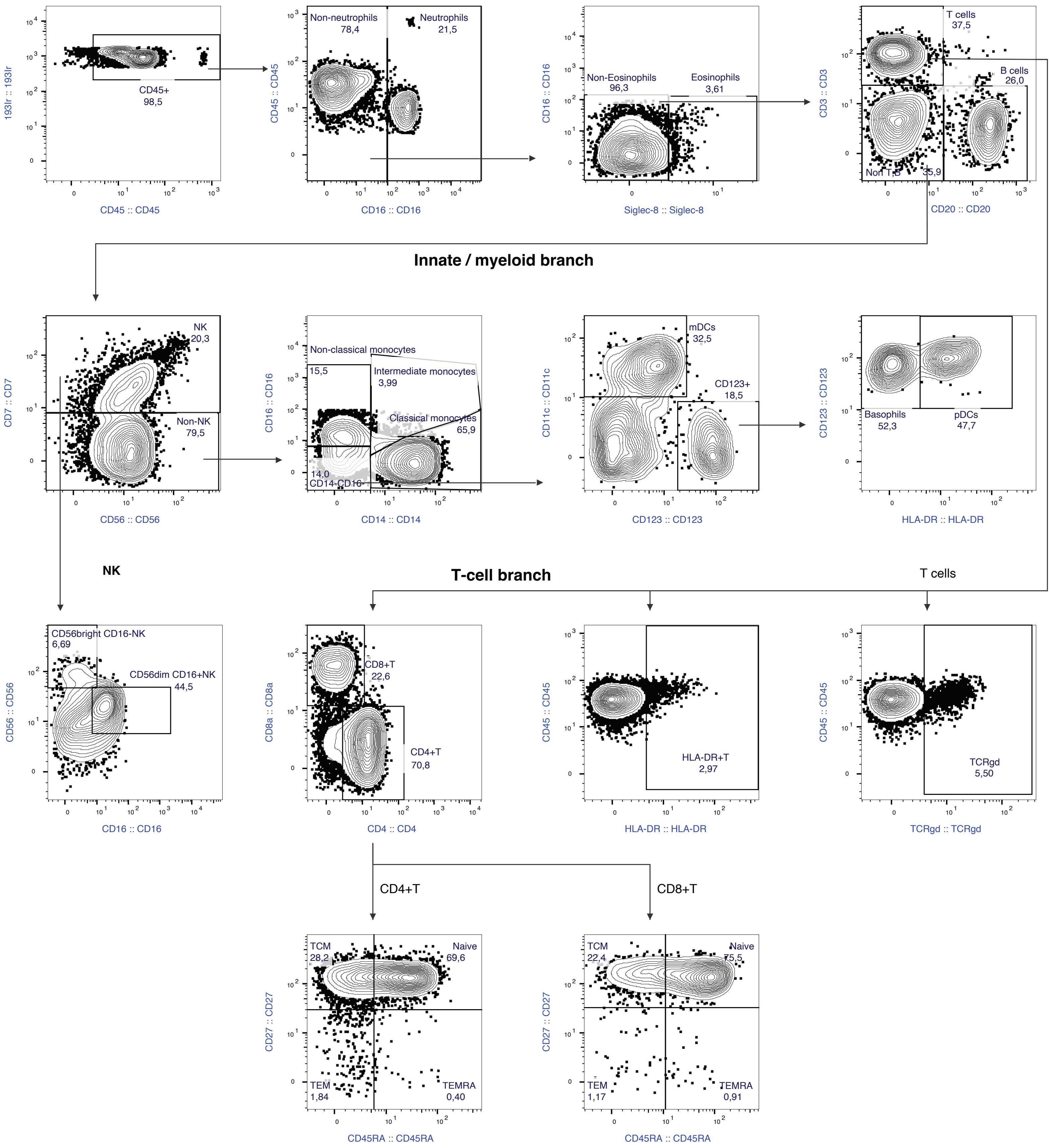


**Supplementary Fig 3. Mass cytometry gating schema.** A manual cell classification schema based on known marker expression patterns.
