## Supplemental Fig 4 for "A Synbiotic Infant Formula Promotes Balanced Immune-Microbiome Interactions in Term Infants: A Randomized Controlled Trial"

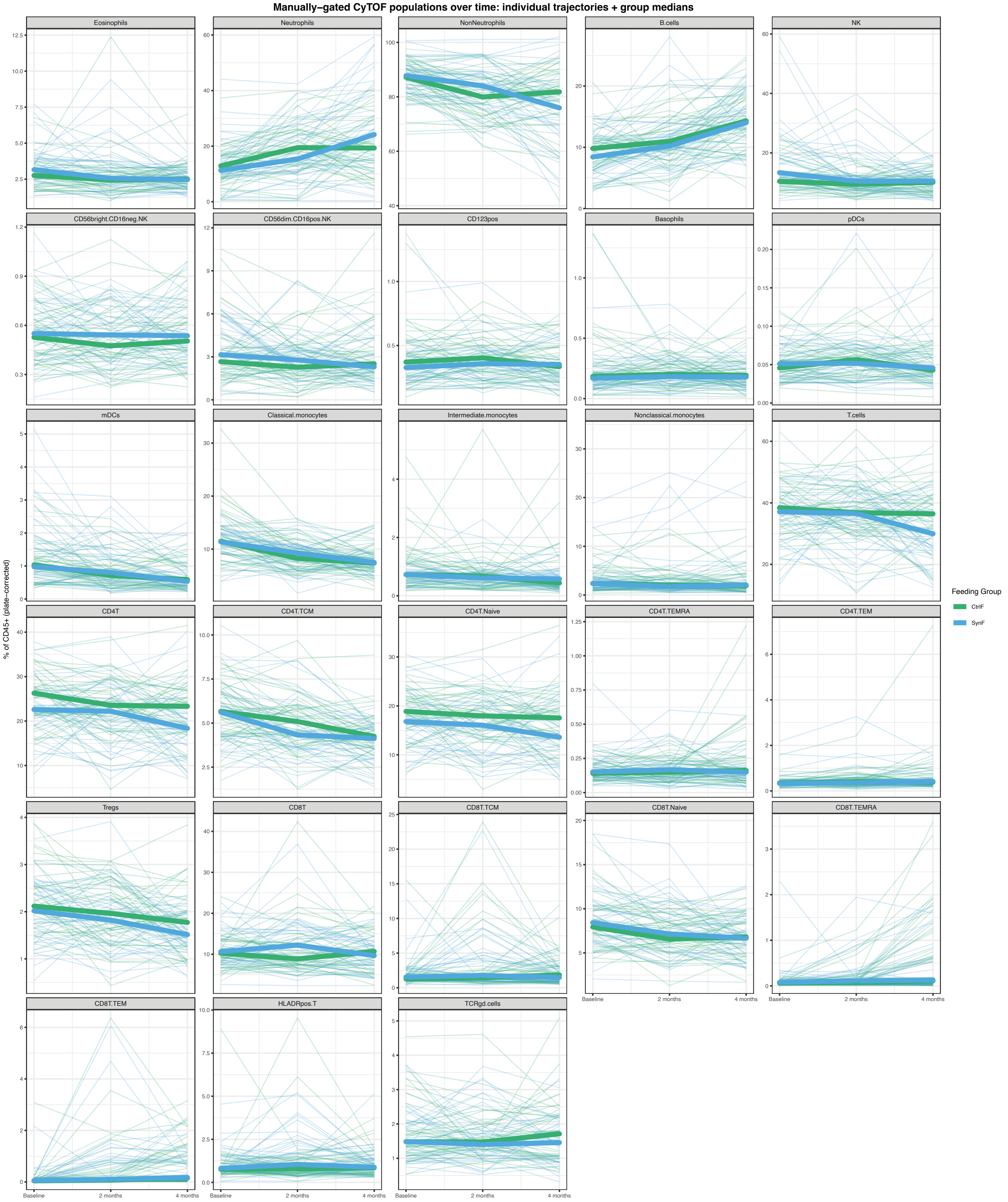


**Supplementary Figure 4. Manually-gated CyTOF cell population frequencies over time, by feeding group**. One panel per manually-gated immune cell population (27 populations, % of $CD45^{+}$ cells, plate-corrected), showing individual-subject trajectories (thin lines) from baseline through 2 and 4 months, colored by feeding group (CtrlF, green; SynF, blue), with group medians overlaid as bold lines with points. Shows the raw longitudinal data underlying the summary statistics reported in Fig. 5a–c and Supplementary Fig. 1–3, for populations across the full compositional range — from abundant lineages (T cells, neutrophils) to rare subsets (basophils, pDCs, $CD123^{+}$ cells) where individual-sample noise is visually apparent.
