## Supplemental Fig 5 for "A Synbiotic Infant Formula Promotes Balanced Immune-Microbiome Interactions in Term Infants: A Randomized Controlled Trial"

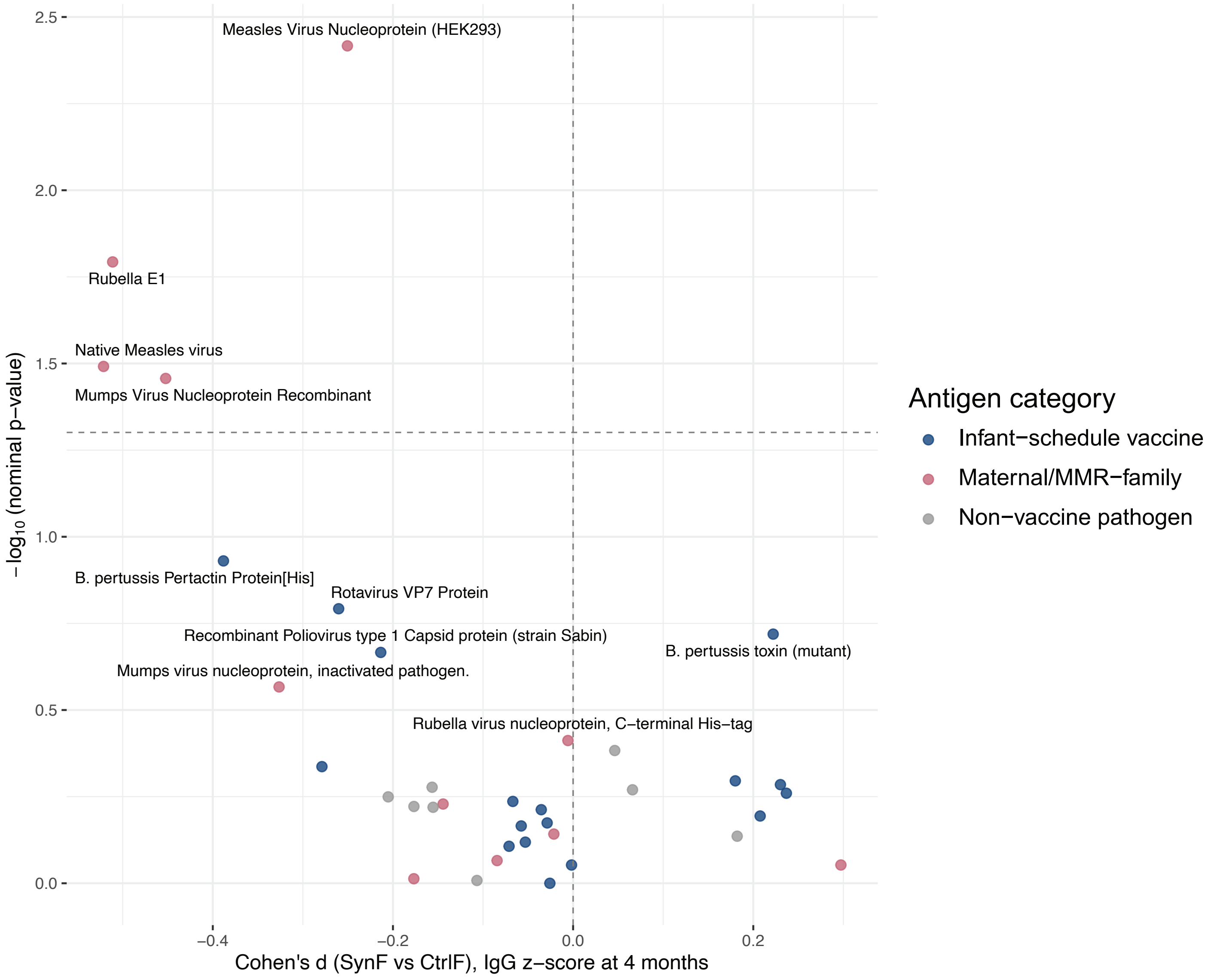


**Supplementary Fig 5. Vaccine antigen measurement at 4 months.** A manual cell classification schema based on known marker expression patterns.
